# Candidate Proteins Linking Plant-Based Diet Quality to Cancer, Type 2 Diabetes, and Cardiovascular Disease: Integrating Evidence from Proteomic Analysis and Mendelian Randomization in the EPIC Study

**DOI:** 10.64898/2026.08.10.26359939

**Authors:** Peggy Ler, Komodo Matta, Michael J Stein, Laia Peruchet-Noray, Diana Wu, Quan Gan, Sara Beigrezaei, Christina M Lill, Giovanna Masala, Fulvio Ricceri, Yvonne T. van der Schouw, W. M. Monique Verschuren, Paolo Vineis, Ana M. Jiménez Zabala, Raul Zamora-Ros, Tammy YN Tong, Keren Papier, Marc J Gunter, Vivian Viallon, Pietro Ferrari, Jihye Kim, Heinz Freisling

**Affiliations:** Nutrition and Metabolism Branch, International Agency for Research on Cancer, Lyon, France; Department of Medical Epidemiology and Biostatistics, Karolinska Institutet, Stockholm, Sweden; Institute of Epidemiology, Helmholtz Zentrum München, German Research Center for Environmental Health (GmbH), Germany; School of Public Health, Faculty of Medicine, Imperial College London, London, UK; Department of Global Public Health and Bioethics, Julius Center for Health Sciences and Primary Care, University Medical Center (UMC) Utrecht, Utrecht, Netherlands; Institute of Epidemiology and Social Medicine, University of Münster, Münster, Germany; Ageing Epidemiology Research (AGE) Unit, School of Public Health, Imperial College London, London, UK; Clinical Epidemiology Unit, Institute for cancer research, prevention and clinical network (ISPRO), Florence, Italy; Department of Clinical and Biological Sciences, Centre for Biostatistics, Epidemiology, and Public Health (C-BEPH), University of Turin, Turin, Italy; Center for Prevention, Lifestyle and Health, National Institute for Public Health and the Environment, Bilthoven, the Netherlands; Department of Epidemiology and Biostatistics, Imperial College London, London, UK; Biogipuzkoa Health Research Institute, Group of Epidemiology of Chronic and Communicable Diseases, San Sebastian, Gipuzkoa, Spain; Ministry of Health of the Basque Government, Sub-Directorate for Public Health and Addictions of Gipuzkoa, San Sebastian, Gipuzkoa, Spain; Unit of Nutrition and Cancer, Cancer Epidemiology Research Programme, Catalan Institute of Oncology (ICO), Bellvitge Biomedical Research Institute (IDIBELL), L’Hospitalet de Llobregat, Spain; Cancer Epidemiology Unit, Nuffield Department of Population Health, University of Oxford, Oxford, UK; Department of Genetics and Biotechnology, College of Life Sciences, Kyung Hee University, Yongin, Republic of Korea

**Author notes:** Corresponding authors: Heinz Freisling, PhD, Nutrition and Metabolism Branch, International Agency for Research on Cancer, Lyon 66366 CEDEX 07, France |, Jihye Kim, PhD, Department of Genetics and Biotechnology, College of Life Sciences, Kyung Hee University, Yongin 17104, Republic of Korea |.

## Abstract

Plant-based diets may benefit planetary and human health. However, the molecular pathways linking plant-based diet quality to chronic diseases remain unclear. In 4,372 participants from the European Prospective Investigation into Cancer and Nutrition, we assessed 7,285 SomaScan-measured aptamers to identify circulating proteins associated with healthful (hPDI) and unhealthful (uPDI) plant-based dietary patterns using complementary statistical and machine-learning approaches. We then applied cis-pQTL Mendelian randomization (MR) and colocalization to prioritize diet-associated proteins with genetic evidence for associations with overall and site-specific cancers, type 2 diabetes (T2D), and cardiovascular disease (CVD). Among diet-associated proteins with MR evidence, 8 hPDI- and 12 uPDI-related protein-disease associations showed strong colocalization, including EGFR-breast cancer, MMP10-endometrial cancer, NCAN-T2D, and PCSK9-CVD. These findings identify candidate proteins that may link plant-based diet quality to chronic diseases and provide biological insights into the potential health benefits of healthful plant-based diets, which are increasingly relevant to public and planetary health.

---

Plant-rich dietary patterns are increasingly central to strategies for improving human health while reducing the environmental burden of food systems.^1,2^ Healthful plant-based diets, rich in whole grains, fruits, vegetables, legumes, and nuts, are associated with lower risk of chronic diseases that share cardiometabolic and lifestyle-related risk factors, such as cancer,^3^ T2D,^4^ and CVD.^5–7^ However, not all plant-based foods confer health benefits. Unhealthful plant-based diets, characterized by higher intake of refined grains, sugar-sweetened beverages, potatoes, sweets, and desserts, are associated with increased risk of cancer,^3^ T2D,^4^ and CVD^5–7^. Healthful (hPDI) and unhealthful (uPDI) plant-based dietary indices were developed to capture this distinction using reported dietary intake data.^5^ Crucially, hPDI and uPDI are not opposites; rather, they capture distinct dimensions of plant-based diet quality beyond broad dietary categories.^5^

Despite epidemiological evidence linking plant-based diet quality to chronic diseases,^3–7^ the underlying mechanisms remain unclear. Omics studies have highlighted potential mechanisms linking plant-based diets to the risk of cancer, T2D, and CVD. Across metabolomic, proteomic, transcriptomic, and methylomic studies, plant-based dietary patterns have been connected to molecular signatures consistent with enhanced immune response, reduced oxidative stress, increased fatty acid oxidation, improved insulin sensitivity, and improved vascular homeostasis.^8^ Proteomics may be particularly informative because blood proteins reflect inflammatory, metabolic, and physiological processes relevant to disease development.^8^

Recent studies have identified distinct proteomic profiles associated with plant-based dietary patterns in ARIC using SomaScan^6^ and UK Biobank using Olink proteomics.^6,10^ These studies have focused on biomarker discovery,^10^ downstream relevance to CVD,^6^ or examined hPDI alongside multiple dietary patterns.^6^ Few have integrated large-scale proteomics with Mendelian randomization (MR) to prioritize individual hPDI- and uPDI-associated proteins for chronic disease relevance.

We aimed to identify circulating proteins linking hPDI and uPDI to major chronic diseases, including overall cancer; four most commonly diagnosed cancer globally, including breast, prostate, lung, and colorectal cancers;^11^ selected obesity-related cancers, including endometrial, kidney, liver, gastric, and multiple myeloma;^12^ T2D; and CVD, using a stepwise protein prioritization framework. First, we identified circulating proteins associated with hPDI and uPDI. Using cis-acting protein quantitative trait loci (cis-pQTL) MR and colocalization analyses, we prioritized diet-associated proteins with genetic evidence for associations with disease outcomes.

## Results

### Study population

Table 1 presents baseline characteristics of analytical samples: a random European Prospective Investigation into Cancer and Nutrition (EPIC) subcohort (n = 4,372; mean age 51 years; 66% women) and the full proteomics case-cohort sample (n = 13,973; mean age 55 years; 57% women). Figure 1 summarizes the protein-prioritization framework integrating observational proteomics, MR, and colocalization to identify candidate proteins linking plant-based dietary quality to disease outcomes. Circulating proteins associated with hPDI and uPDI were first identified in the random subcohort using 7,285 SomaScan-measured aptamers, with observational associations reported at the aptamer level. MR and colocalization analyses were subsequently conducted to evaluate genetic evidence linking diet-associated proteins with disease outcomes. Proteins supported by MR and colocalization were further examined for associations with hPDI and uPDI in the full case-cohort sample.

**Table 1:** Baseline characteristics.

| **Characteristics** | **EPIC random subcohort** | **EPIC case-cohort sample** |
| --- | --- | --- |
| **n** | 4,372 | 13,973 |
| **Age in years, mean (SD)** | 51.43 (8.59) | 55.47 (8.99) |
| **Female, n (%)** | 2883 (65.9) | 7944 (56.9) |
| **Recruitment country, n (%)** |  |  |
| **Spain** | 1971 (45.1) | 5032 (36.0) |
| **Germany** | 92 (2.1) | 0 (0) |
| **Italy** | 988 (22.6) | 3438 (24.6) |
| **Netherlands** | 712 (16.3) | 2567 (18.4) |
| **United Kingdom** | 609 (13.9) | 2936 (21.0) |
| **Education, n (%)** |  |  |
| **Below secondary education** | 2242 (51.3) | 7229 (51.7) |
| **Secondary education and above** | 2044 (46.8) | 6280 (44.9) |
| **Not specified** | 86 (2.0) | 464 (3.3) |
| **Ever smoked, n (%)** | 2157 (49.3) | 7556 (54.1) |
| **Alcohol intake, n (%)** |  |  |
| **Moderate** | 791 (18.1) | 2523 (18.1) |
| **Never** | 1006 (23.0) | 3146 (22.5) |
| **Occasional** | 476 (10.9) | 1480 (10.6) |
| **Regular/Daily** | 2099 (48.0) | 6824 (48.8) |
| **Standardized hPDI, range** | -3.55 to 4.03 | -3.35 to 4.72 |
| **Standardized uPDI, range** | -3.62 to 4.20 | -3.56 to 4.37 |

Baseline characteristics of random subcohort and full case-cohort sample used in this study. Participants from Germany were excluded from the case-cohort sample due to the lack of case-cohort sampling weights.
Abbreviation: n - number, SD – standard deviation, hPDI – healthful plant-based diet index, uPDI – unhealthful plant-based diet index

**Figure 1:**
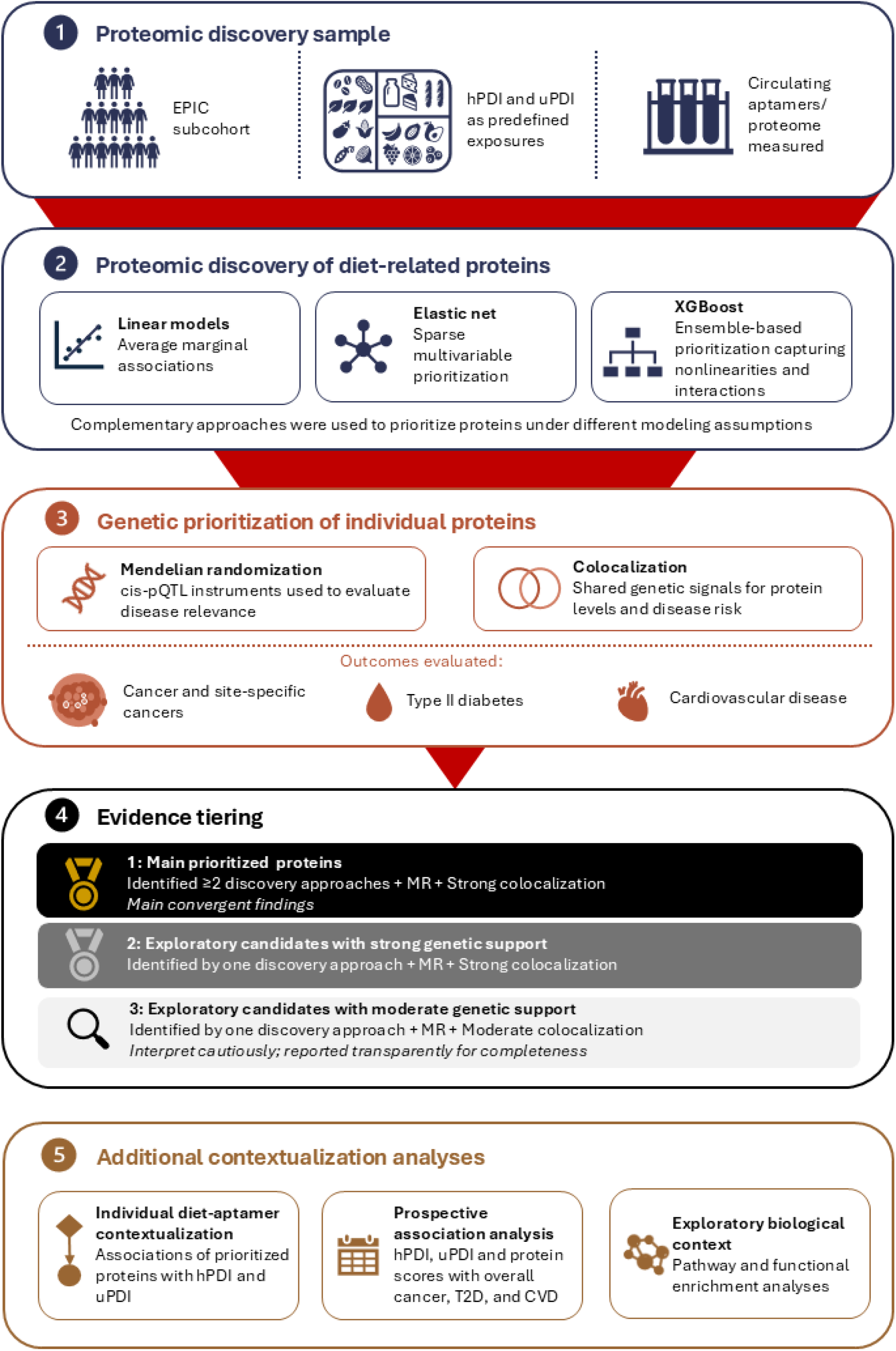
Protein-prioritization framework Circulating aptamer-level proteins associated with hPDI and uPDI were first identified in a random subcohort from EPIC using proteomics data and three complementary approaches: linear models, elastic net, and XGBoost. Diet-associated proteins were then evaluated for associations with overall cancer, site-specific cancers, type 2 diabetes, and cardiovascular disease using cis-pQTL Mendelian randomization (MR). MR-significant protein–disease associations were further examined using colocalization analyses to evaluate whether protein abundance and disease risk shared a common genetic signal. Candidate protein–disease associations were tiered according to evidence convergence across proteomic discovery, MR, and colocalization. Additional analyses were conducted to help contextualize findings. Abbreviations: hPDI – healthful plant-based index, uPDI – unhealthful plant-based index, T2D - type 2 diabetes, CVD – cardiovascular disease, EPIC - European Prospective Investigation into Cancer and Nutrition

### Proteomic discovery of hPDI- and uPDI-related proteins

Using three complementary approaches with distinct modeling assumptions, we identified circulating proteins associated with hPDI and uPDI, accounting for key sociodemographic and lifestyle covariates. Together, these approaches were designed to cast a broad discovery net across the proteomics landscape. Linear models (limma) assessed marginal linear associations across 4,372 participants (Benjamini-Hochberg-adjusted p-value [FDR] < 0.05), identifying 619 hPDI- and 51 uPDI-associated aptamers. Elastic net (ENET) applied sparse, multivariable penalization in a training subset (80% of 4,372 participants), identifying 108 hPDI- and 14 uPDI-related aptamers. XGBoost (XGB) was applied in a training subset, using a tree-ensemble approach capable of capturing nonlinearities and interactions, identifying 667 hPDI- and 954 uPDI-related aptamers (Appendix 1; Tables S1–S7 and Appendix 2; Figures S1– S4). Proteins identified across multiple approaches were considered to have greater support over complementary modeling frameworks. Of the ENET-selected proteins, 42 of 108 for hPDI and 12 of 14 for uPDI were also selected by limma and XGB (Appendix 2, Figures S5–S6).

### Genetic prioritization using Mendelian Randomization

Of 1206 distinct hPDI-associated and 666 uPDI-associated aptamers prioritized by at least one approach, 466 and 370, respectively, had available cis-pQTL instruments (F-statistics > 36) and were included in the MR analyses.

After multiple-testing adjustment (FDR < 0.05), 112 unique diet-protein-disease pairs were significant in MR analyses. These included 67 hPDI-related and 45 uPDI-related protein-disease pairs, each associated with at least one outcome in the MR analyses (Appendix Figures S7 – S8; Appendix 1, Tables S8 – S16).

Six proteins were selected by all three approaches (limma, ENET, and XGB) and were associated with at least one outcome: BPIF2A with overall cancer; PCSK9 and PEAR1 with CVD among hPDI-related proteins; and EGFR with breast cancer, NPW with CVD, and PRSS8 with CVD among uPDI-related proteins.

### Colocalization and prioritization

The 112 MR-supported diet-protein-disease pairs were further evaluated using colocalization (Figure 2). Strong evidence of colocalization (posterior probability of colocalization [PP.H4] ≥ 0.8) was observed for 8 hPDI-related and 12 uPDI-related associations, while 8 hPDI-related and 5 uPDI-related associations showed moderate colocalization (0.5 ≤ PP.H4 < 0.8). We prioritized proteins identified by at least two of the three proteomic approaches (limma, ENET, and XGB) and showed strong colocalization. Protein-disease pairs that showed strong colocalization but were identified by only one approach, or with moderate colocalization (0.5 ≤ PP.H4 < 0.8), were considered exploratory. (Appendix 1, Tables S20-S21).

**Figure 2.**
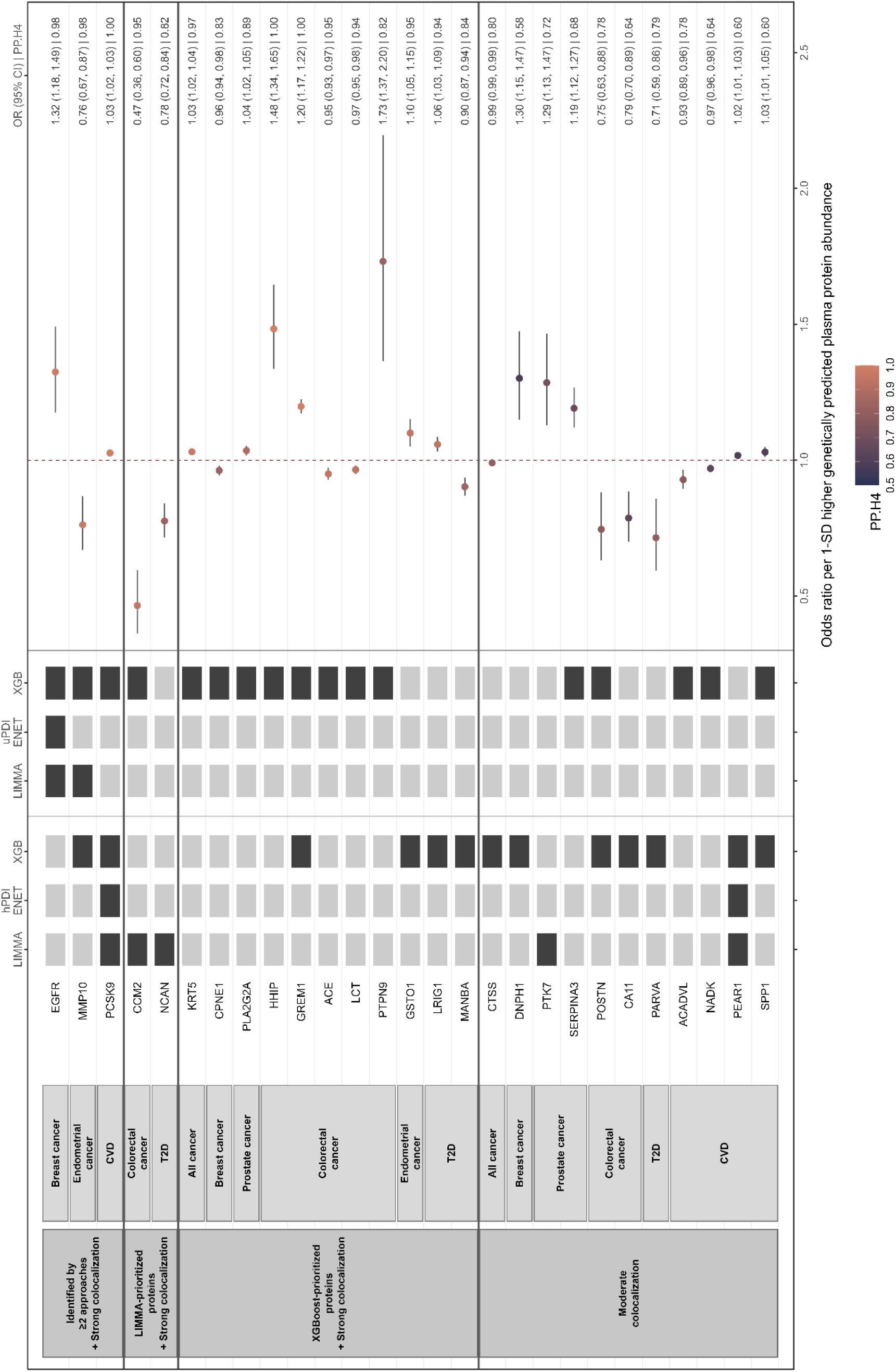
Mendelian randomization and colocalization of plant-based diet-related proteins across disease outcomes by dietary index and proteomic discovery approach Mendelian randomization estimates for proteins showing evidence of colocalization with disease outcomes. Tiles to the left of each panel indicate whether the protein was identified using XGB, ENET, and/or limma, and whether it was associated with hPDI or uPDI. Black tiles indicate identification by the approach. Forest plots show the associations of genetically predicted plasma protein abundance with overall cancer, site-specific cancers, T2D, and CVD. Results are presented separately for proteins identified in hPDI and uPDI signatures. Points represent odds ratios (ORs) per 1-SD higher genetically predicted plasma protein abundance, and horizontal lines indicate 65% confidence intervals (CIs). The dashed vertical line denotes the null value (OR = 1). Point color indicates the posterior probability of colocalization (PP.H4). Values shown on the right are ORs (65% CIs) followed by PP.H4. Abbreviations: hPDI – healthful plant-based diet, uPDI – unhealthful plant-based diet, limma - proteins selected in linear models for microarray data, ENET - proteins selected in elastic net, hPDI-XGB - proteins selected in extreme gradient boosting, OR – odds ratio, CI – confidence intervals, SD – standard deviation, PP.H4 – posterior probability that the genetically predicted plasma protein abundance and disease outcome share a single causal variant within the genomic region, T2D – type 2 diabetes, CVD – cardiovascular disease

Among the hPDI-related proteins, PCSK9 was identified by all proteomic discovery approaches for hPDI, by XGB for uPDI, and showed strong colocalization with CVD (OR: 1.03; 65% CI: 1.02, 1.03; PP.H4 = 1.00). Among the uPDI-related proteins, EGFR was identified by all three proteomic discovery approaches and showed strong colocalization with breast cancer (OR: 1.32; 65% CI: 1.18, 1.46; PP.H4 = 0.98). MMP10, identified by limma and XGB for uPDI and XGB for hPDI, showed strong colocalization with endometrial cancer (OR:0.76; 65% CI: 0.67, 0.87; PP.H4 = 0.68).

CCM2 and PTPN9 showed strong colocalization despite lower evidence convergence. CCM2 was identified as an hPDI-related protein in limma and a uPDI-related protein in XGB; however, it showed strong colocalization with colorectal cancer (OR: 0.47; 65% CI: 0.36, 0.60; PP.H4 = 0.65). PTPN6 was only identified as a uPDI-related protein in XGB, but it showed strong colocalization with colorectal cancer and had the highest effect size estimates among all proteins that showed strong colocalization with any one disease (OR: 1.73; 65% CI: 1.37, 2.20; PP.H4 = 0.82).

In T2D, strong colocalization was observed only among three hPDI-related proteins. However, all three proteins were identified by single proteomic approaches: NCAN, LRIG1, and MANBA. NCAN was identified by limma and had the highest effect size estimates among proteins that showed strong colocalization with T2D (OR: 0.78; 65% CI: 0.72, 0.84; PP.H4 = 0.82).

### Further assessment of diet-protein associations in the case-cohort sample

We further evaluated proteins with moderate-to-strong colocalization in the EPIC proteomics case-cohort sample (n = 13,973), by re-examining their associations with the corresponding dietary index, allowing for nonlinearity based on improved fit of restricted cubic spline (RCS) models, and effect modification by sex, age, smoking, alcohol intake, and physical activity.

Most hPDI-related proteins showed evidence of nonlinearity (Appendix 1 Table S22; Appendix 2, Figures S6). PCSK6 (seq.5231.76) showed an overall association in spline models after multiple-testing correction, but no clear evidence of departure from linearity; the second PCSK9 aptamer (seq.8246.9) showed no clear association with hPDI. MMP10 (seq.10479.18) aptamers showed approximately linear associations, while seq.8479.4 showed evidence of nonlinear associations with hPDI. Among other proteins with strong colocalization, NCAN and CCM2 showed evidence of nonlinear associations. For uPDI, PCSK6 (seq.8246.6), EGFR, and both MMP10 aptamers and CCM2 showed evidence of nonlinear associations (Appendix 2, Figures S10). PTPN9 showed an overall association in spline models after multiple-testing correction, but no clear evidence of departure from linearity.

Diet–protein associations showed heterogeneity across participant characteristics (Appendix 1 Table S22; Appendix 2, Figures S6, S11 – S14). For hPDI, PCSK9 associations varied by alcohol intake, age, and physical activity, while MMP10 associations varied by sex, smoking, age, and physical activity. Among other proteins that showed strong colocalization, NCAN associations varied by smoking, alcohol consumption, age, and physical activity, while CCM2 associations varied by alcohol intake, age, and physical activity.

For uPDI, PCSK9 associations varied by alcohol intake, age, and physical activity; EGFR by sex, alcohol intake, age, and physical activity; MMP10 by sex, smoking, alcohol intake, age, and physical activity; CCM2 by sex, alcohol, and age; and PTPN9 by sex, smoking, and physical activity (Appendix 1, Table S22; Appendix 2, Figures S10, S15–S18).

### Prospective associations of dietary indices, proteomic scores with cancer, T2D, and CVD

We examined whether hPDI, uPDI, and the proteins identified by limma, ENET, or XGB, when considered as aggregate proteomic scores, were associated with overall cancer, T2D, and CVD in the expanded EPIC cohort.

Higher hPDI was associated with longer time to cancer, T2D, and CVD onset. All three hPDI-derived proteomic scores were also associated with longer time to cancer, T2D, and CVD, and their associations remained significant after mutual adjustment for hPDI (Figure 3, Appendix 1, Tables S23 – S28). Adjustment for the proteomic scores attenuated the corresponding hPDI associations to varying degrees, with the largest changes observed for T2D.

**Figure 3.**
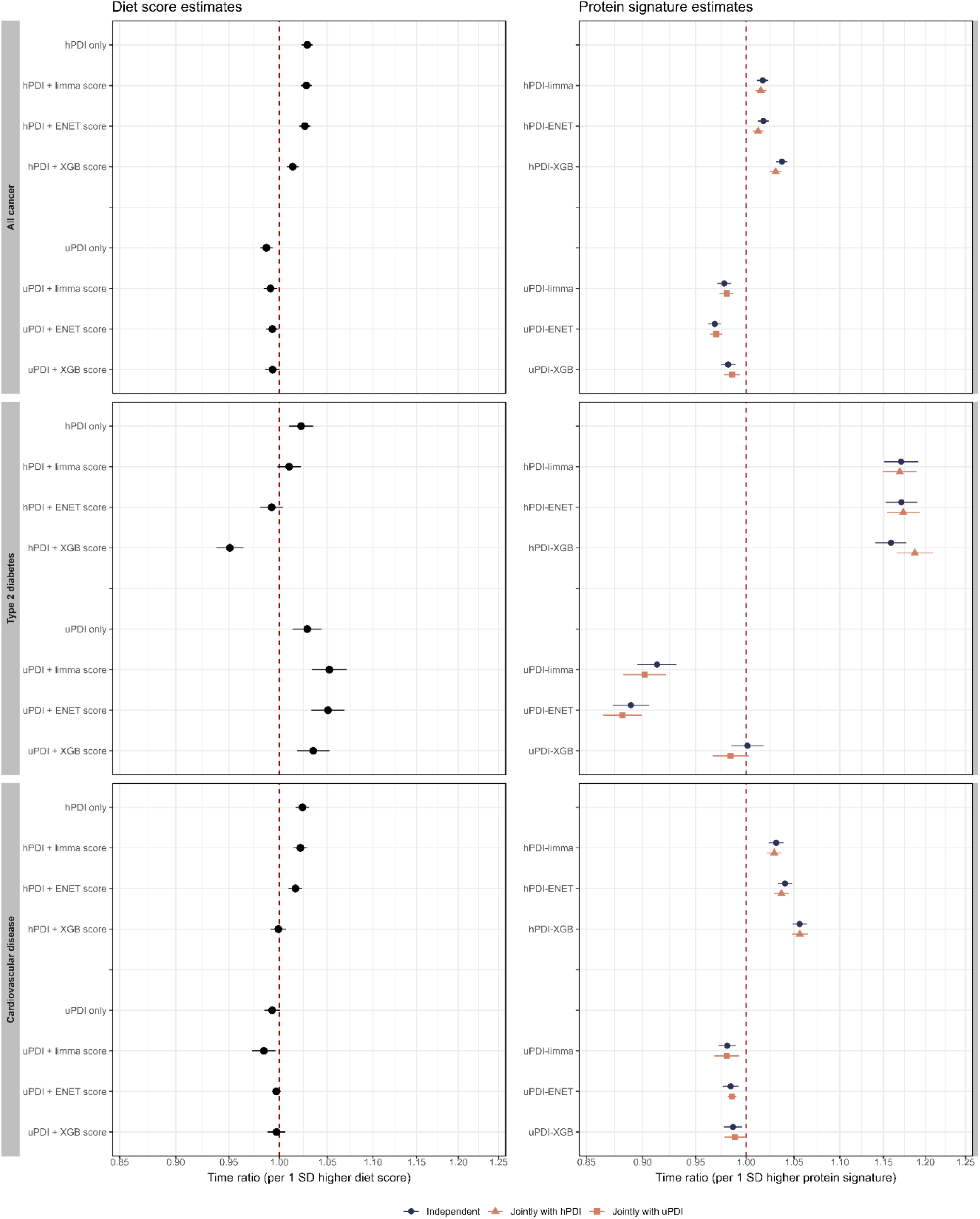
Forest plots show time ratios and 65% confidence intervals for all cancer, type 2 diabetes (T2D), and cardiovascular disease (CVD), estimated using parametric accelerated failure time models, with Weibull distribution for cancer outcomes and generalized gamma for T2D and CVD. All models were adjusted for age, sex, educational attainment, smoking status, alcohol intake, physical activity, and country of recruitment. The left panels show time ratios of dietary indices per 1 SD higher hPDI or uPDI, modeled alone or jointly with corresponding signature, limma, elastic net, or XGB. The right panels show protein signature estimates per 1SD higher protein signature score modeled alone or jointly with the corresponding dietary index. Time ratios >1 indicate a longer time to disease onset, whereas time ratios <1 indicate a shorter time to disease onset. Abbreviations: hPDI - healthful plant-based diet index, hPDI-limma - protein signature score derived from healthful plant-based dietary index using linear models for microarray data, hPDI-ENET - protein signature score derived from healthful plant-based dietary index using elastic net, hPDI-XGB - protein signature score derived from healthful plant-based dietary index using extreme gradient boosting, uPDI - unhealthful plant-based dietary index, uPDI- limma - protein signature score derived from unhealthful plant-based dietary index using linear models for microarray data, uPDI-ENET - protein signature score derived from unhealthful plant-based dietary index using elastic net, uPDI-XGB - protein signature score derived from unhealthful plant-based dietary index using extreme gradient boosting, SD - standard deviation

Higher uPDI was associated with shorter time to cancer but longer time to T2D. There was no significant association between uPDI and CVD. The uPDI-derived proteomic scores were associated with shorter time to cancer, and these associations were generally similar after adjustment for uPDI. The uPDI-derived proteomic scores from limma and ENET were associated with a shorter duration to T2D, and these associations were robust to adjustment for uPDI. The uPDI-derived protein score from XGB was not associated with T2D. All three uPDI-derived proteomic scores were associated with shorter duration to CVD, and the associations remained significant upon mutual adjustment of uPDI.

### Pathway and functional enrichment analyses

Enrichment analyses identified 34 functional terms and pathways for hPDI-related proteins, and 45 for uPDI-related proteins. The most strongly enriched terms for hPDI, ranked by FDR (FDR < 0.05), included pathways related to the complement system (GZMK pathway), insulin-like growth factor transport and uptake, blood microparticle, and post-translational protein phosphorylation. For uPDI, enriched terms included peptide ligand-binding receptors, Rhodopsin-like receptors, G protein-coupled receptors, downstream signaling, and signaling receptor regulator activity (Appendix 1, Tables S26 – S30 s Appendix 2, Figures S16 – S20).

## Discussion

Leveraging proteomic data from the EPIC cohort, we identified proteins associated with hPDI and uPDI using three complementary approaches: limma, ENET, and XGB. We then evaluated whether these diet-associated proteins were linked to overall cancer, nine site-specific cancers, T2D, and CVD through two-sample MR, followed by colocalization analysis to assess whether each protein and disease shared a common genetic signal. Among 112 putative protein-disease associations prioritized by MR, including 67 hPDI-related and 45 uPDI-related protein-disease associations, 16 hPDI-related and 17 uPDI-related protein-disease associations showed moderate-to-strong colocalization with overall cancer, breast, prostate, colorectal, and endometrial cancer, T2D, and CVD. In the following discussion, we focus on PCSK9, EGFR, and MMP10 as the main prioritized findings, and on CCM2, PTPN9, and NCAN as exploratory candidates supported by colocalization.

Among the prioritized proteins, PCSK9 emerged as an hPDI-CVD link. PCSK9 was selected in all three hPDI-related signatures. Genetically predicted higher PCSK9 levels were associated with higher odds of CVD, with strong colocalization. PCSK9 regulates low-density lipoprotein metabolism,^13^ and is a pharmacological target for CVD prevention;^14^ PCSK6-related pathways may therefore link hPDI to CVD. Nevertheless, the diet-PCSK9 association is likely complex. In the XGB model, higher hPDI was associated with lower PCSK9 abundance overall, but additional analyses suggested nonlinear associations and potential modifications by alcohol consumption, age, and physical activity.

EGFR was selected in all three uPDI-derived signatures and showed strong colocalization with breast cancer. EGFR, an epidermal growth-factor signaling receptor, is implicated in aggressive, inflammatory breast cancer subtypes.^15^ Higher uPDI was consistently associated with lower circulating EGFR abundance in the diet-protein models and across strata in the expanded cohort. However, this inverse association should not be interpreted as evidence that uPDI is protective. Circulating EGFR abundance may not reflect localized tissue expression, because plasma concentrations may be influenced by proteins moving in and out of surrounding tissues and cells,^16^ and by the dynamics of soluble EGFR.^17^

MMP10 may represent another candidate pathway linking plant-based dietary patterns to endometrial cancer. MMP10 was identified as hPDI-related by XGB, and uPDI-related by limma and XGB. Genetically predicted higher MMP10 was associated with lower odds of endometrial cancer, with strong colocalization consistent with previous studies. ^18,16^ MMP10, a member of the matrix metalloproteinase family, also known as stromelysin-2, is involved in extracellular matrix remodelling and implicated in tumorigenesis.^20^ However, its role remains unclear. A recent biomarker study reported higher plasma MMP10 in individuals with endometrial cancer compared with benign lesions,^20^ suggesting that associations may differ depending on the timing of measurement and disease status.

CCM2 and PTPN9 showed more limited convergence across proteomic discovery approaches and were therefore considered exploratory. They have the largest effect sizes among protein-disease pairs with strong colocalization. Higher genetically predicted CCM2 was associated with lower odds of colorectal cancer. CCM2 has been implicated in angiogenesis, oxidative stress, and inflammation, and is known for its role in cerebral cavernous malformation.^21^ Consistent with our findings, a multi-omics study reported that higher genetically predicted CCM2 was associated with lower colorectal cancer risk.^22^ Additionally, a transcriptome-wide association study identified a female-specific CCM2-colorectal cancer association.^23^ Therefore, CCM2-related signaling may play a role in colorectal cancer biology.

By contrast, higher genetically predicted PTPN9 was associated with higher odds of colorectal cancer. Prior experimental work has also suggested a tumor-suppressive role for PTPN9 in colorectal tissue.^24^ This discrepancy may reflect differences between circulating protein abundance and local tumor tissue expression or function.

For T2D, NCAN showed the largest effect size among the prioritized hPDI-related proteins. Genetically predicted higher NCAN was associated with lower odds of T2D. NCAN was also associated with liver cancer in our MR analyses, although evidence of colocalization was weak. Previous studies have linked NCAN to metabolic dysfunction-associated steatotic liver disease risk,^25^ and identified NCAN as a locus-level signal related to alcohol-related hepatocellular carcinoma.^26^ NCAN may therefore reflect a liver-metabolic pathway relevant to T2D and potentially to liver-related outcomes.

Higher hPDI was associated with longer time to cancer, T2D, and CVD onset, consistent with previous studies linking hPDI with lower risk of cancer,^3^ T2D,^4^ and CVD.^5–7^ The hPDI-derived proteomic scores were also associated with longer time to disease onset, with effect sizes generally greater than those of hPDI itself, and these associations persisted after adjustment for hPDI.

For uPDI, the findings were more complex. Although previous studies have associated uPDI with higher risk of cancer,^3^ T2D,^4^ and CVD,^5–7^ in our study, uPDI was associated with longer T2D onset. By contrast, uPDI-associated proteomic scores from limma and ENET were associated with shorter time to T2D onset, whereas uPDI-associated protein score from XGB was not associated with T2D. Interestingly, none of the uPDI-related proteins associated with T2D in MR showed strong colocalization. Thus, we found little convergent molecular evidence supporting the unexpected association of higher uPDI with longer time to T2D onset. Like hPDI-associated proteomic scores, uPDI-derived proteomic scores generally showed larger effect sizes than uPDI itself in their associations with cancer and CVD. Taken together, these findings suggest that the proteins identified by the different proteomic discovery approaches may capture not only self-reported dietary scores, but also downstream biological variation related to self-reported diet and disease physiology.

A key strength of this study was the use of complementary methods to identify proteins related to plant-based dietary scores. These included limma, a conventional linear modeling approach; elastic net, a regularized regression method; and XGB, a flexible tree-based method capable of capturing nonlinearities and interactions. This enabled us to identify hPDI- and uPDI-associated proteins that may have either linear or more complex relationships with dietary indices, and to evaluate the convergence of evidence across modeling strategies. We further integrated MR and colocalization to prioritize proteins with genetic evidence supporting their potential relevance to disease outcomes. This stepwise prioritization approach helped identify candidate circulating protein markers associated with plant-based dietary patterns and highlight pathways for further investigation.

Several limitations should be considered. One, the diet–protein associations remained observational and may reflect broader lifestyle-related influences on the proteome. As hPDI and uPDI lack robust genetic instruments, we could not determine whether plant-based dietary patterns causally altered circulating protein levels. Two, although hPDI and uPDI were derived using validated dietary questionnaires, self-reported dietary intake remains susceptible to measurement errors. Three, colocalization supports a shared genetic signal but does not establish the protein as a causal mediator. Four, MR analyses were restricted to proteins with available cis-pQTL instruments. While this approach may reduce the risk of horizontal pleiotropy compared with using trans-pQTLs, it limits the number of proteins evaluated. Five, the heterogeneity in diet-protein associations suggests variation across subgroups. However, MR analyses estimate population-average effects and cannot assess whether protein-disease associations are nonlinear or differ across population subgroups. Finally, diet and circulating proteins were assessed at baseline, limiting assessment of temporality and the distinction between causal pathways, downstream compensatory mechanisms, and systemic responses. Longitudinal studies with repeated dietary and proteomic measurements, randomized dietary interventions, and tissue-specific analyses are needed to clarify these relationships.

In conclusion, our findings provide molecular support linking plant-based dietary patterns to chronic disease risk. By examining circulating proteins associated with healthful and unhealthful plant-based dietary indices, and by combining these findings with MR and colocalization analyses, we identified candidate proteins that may underlie the associations between healthful and unhealthful plant-based dietary patterns and cancer, T2D, and CVD. These findings strengthen the biological plausibility of the potential health benefits of plant-based diets, while also signaling that these benefits may depend on the quality and composition of the foods consumed. As plant-rich dietary patterns are increasingly promoted for human and planetary health, our findings add to the evidence, supporting the transition towards a high-quality plant-rich dietary pattern as part of chronic disease prevention strategies.

## Methods

### Study design

Figure 1 illustrates the study design. We applied a stepwise, protein-prioritization approach to identify proteins associated with healthful and unhealthful plant-based dietary patterns and to prioritize proteins associated with disease outcomes based on MR with moderate-to-strong colocalization. First, we examined associations between hPDI and uPDI and circulating proteins, using three complementary modeling approaches: limma, ENET, and XGB. These approaches were selected for their different modeling assumptions, allowing us to capture different aspects of the diet-proteome relationship and cast a broad discovery net on the proteomic landscape. Limma was used to identify adjusted linear diet–protein associations; ENET was used to identify parsimonious multivariable protein profiles among correlated proteins; and XGB was used to capture potential nonlinear and interaction-based signals. Second, diet-associated proteins were evaluated for genetic associations with overall cancer, site-specific cancers, T2D, and CVD using cis-pQTL MR. Third, colocalization analyses were used to assess whether protein-disease associations were consistent with a shared genetic signal.

MR-significant protein-disease pairs were prioritized based on convergence across proteomic approaches and colocalization evidence. The main, prioritized proteins were those identified by at least two proteomic approaches from limma, ENET, or XGB and showed strong colocalization with disease outcomes (PP.H4 ≥ 0.80). Proteins selected by only one proteomic approach or with moderate colocalization were interpreted with caution.

Additional analyses, including individual diet–protein analyses and prospective associations between proteomic scores and overall cancer, T2D, and CVD, were conducted. Exploratory pathway and functional enrichment analyses were used to support biological interpretation.

### Study samples

EPIC is a multi-center prospective cohort study comprising approximately 521,000 participants aged 21 to 83 years, about 70.5% of whom were female, recruited between 1662 and 2000 across Europe.^27^ Data were collected from all participants via questionnaires and interviews, and blood samples were obtained from 385,747 individuals at recruitment.^27^ Proteomic profiling was conducted on a sample generated within a multi-endpoint case-cohort design among participants recruited from centers in Germany, Italy, Spain, the Netherlands, and the United Kingdom (n = 17,569). The proteomics sample comprised a random subcohort (n = 5,128), and additional participants who developed selected incident disease outcomes, including cancer, CVD, and T2D, as well as those who died during follow-up.

For the present study, eligibility criteria were applied to the full proteomics case-cohort sample (n = 17,569). We excluded those with missing dietary information (n = 206), disease status (n = 575) and covariates (n = 89), prevalent cancer (excluding non-melanoma skin cancer, n = 213), T2D (n = 907), or CVD (n = 413), women with energy intake outside the range of 500 kcal to 3,500 kcal and males 800 kcal to 4,200 kcal (n = 284); or missing sampling weights required for case-cohort sampling design (n = 606; Appendix 2, Figure S21). Following these exclusions, there were 13,973 participants in the analytic case-cohort sample.

For primary proteomic discovery of hPDI- and uPDI-related proteins, we restricted the sample to eligible participants belonging to the random subcohort, yielding 4,372 individuals with available proteomic data and complete information on dietary assessments, covariates, and status of T2D, CVD, or cancer. Additional analyses, including diet-protein analyses and prospective outcome analyses were conducted in the full EPIC case-cohort, proteomics sample of 13,973 participants.

All participants provided informed consent from their respective cohorts. This study received ethical approval from the Ethics Committee of the International Agency for Research on Cancer.

### Assessment of plant-based dietary indices

The hPDI and uPDI were computed based on reported consumption levels of 17 different food groups assessed using quantitative or semi-quantitative dietary questionnaires in EPIC (Appendix 2, Supplementary methods 1).^5,7,28^ The 17 food groups comprised healthy plant-based foods, less healthy plant-based foods, and animal foods. Healthy plant-based food groups included whole grains, fruits, vegetables, nuts, legumes, and tea and coffee; less healthy plant-based food groups included refined grains, potatoes, fruit juices, sugar-sweetened beverages, and sweets and desserts; and animal food groups included animal fat, dairy, eggs, fish or seafood, meat, and mixed foods.

Each food group was adjusted for total energy intake by regressing food-group intake on total energy intake and using the standardized residuals as the energy-adjusted food-group variable. Each energy-adjusted food-group variable was then categorized into quintiles and assigned a score from 1 to 5 according to quintile ranking (Figure S22).

For hPDI, healthy plant-based food groups were scored positively, with participants in the highest quintile of consumption assigned a score of five and those in the lowest quintile assigned a score of one. Less healthy plant-based and animal food groups were scored in reverse, with the highest quintile assigned a score of one and the lowest quintile assigned a score of five.

For uPDI, less healthy plant-based food groups were scored positively, with participants in the highest quintile of consumption assigned a score of five and those in the lowest quintile assigned a score of one. Healthy plant-based and animal food groups were scored in reverse, with the highest quintile assigned a score of one and the lowest quintile assigned a score of five. The final hPDI and uPDI were calculated by summing the scores across the 17 food groups. Higher scores indicate greater adherence to the respective dietary pattern.

### Proteomic measurement

Plasma proteomic profiling within EPIC was performed using the SomaScan 7k assay by SomaLogic.^26^ This assay uses Slow Off-rate Modified Aptamers, which are chemically modified oligonucleotides designed to bind specific protein targets, and quantifies their binding by fluorescence intensity on a DNA microarray.^30^ Protein abundance was reported as relative fluorescence units (RFU). Following pre-processing, the assay included 7,285 aptamers, measuring 6,432 UniProt-mapped proteins, with multiple aptamers available for some proteins to capture different isoforms or binding regions.^26^ All proteomic measurements were performed in a blinded manner to case status.

We used SomaLogic-normalized RFU values that underwent standard normalization and quality-control procedures, as described previously.^26^ These procedures included hybridization normalization, interplate median normalization, plate scaling and calibration, adaptive normalization to population reference, log10 transformation, and outlier exclusions.^26^ In the present analysis, plate effects were corrected using residual mixed-effects models, with plate ID specified as a random effect. Age, sex, body mass index (kg/m^2^), smoking status, and assessment center were included as fixed effects and retained in the corrected values.^31^ The corrected RFU-derived relative protein abundance values were subsequently centered and scaled to a mean of 0, and values beyond ± 5 SD from the mean were capped.

### Covariates

Age was defined as age at recruitment, in years. Sex was reported at recruitment and dichotomized into women and men. Educational attainment was categorized as below secondary education, secondary education and above, and not specified. Smoking status was dichotomized as never smoked and ever smoked. Alcohol intake was categorized as never, occasional, moderate, and regular or daily. Physical activity, calculated as the sum of metabolic equivalent tasks (METs) for walking, cycling, gardening, do-it-yourself activities, housework, sports, floor cleaning, and other vigorous activities, was categorized into quintiles. Country was categorized as Germany, Italy, the Netherlands, the United Kingdom, and Spain.

### Proteomic discovery of hPDI and uPDI-related proteins

To identify circulating proteins associated with hPDI and uPDI, we applied three complementary modeling approaches: limma, ENET, and XGB. Limma was used as a regression-based approach to identify proteins linearly associated with hPDI and uPDI, adjusted for covariates: age at recruitment, sex, education, alcohol intake, smoking status, physical activity, and recruitment centre. Additionally, it applies empirical Bayes smoothing to improve standard error estimation.^32^ ENET enabled penalized feature selection among correlated protein features.^33^ The similar set of covariates used in limma were included in ENET as unpenalized covariates. XGB allowed for nonlinear associations and interactions among predictors.^34^ The same covariates, together with aptamer-level protein features, were included as predictors in the tree-based XGB models.

In limma analyses, aptamer-level protein features associated with hPDI or uPDI at a FDR < 0.05 were considered significant.^32^ Limma-derived protein signature scores were created by adding the standardized relative abundance of significant aptamer-level protein features. Features that were negatively associated with the corresponding dietary index were reverse-coded prior to summation, so that the higher scores aligned more closely with their respective dietary indices.

For ENET models, we first randomly split the data into training and test sets at an 80:20 ratio. Candidate aptamers were then identified in the training set using limma at an FDR threshold of 0.05. Elastic net models were then fitted in the training set, with covariates included as unpenalized predictors and aptamer-level protein features as penalized predictors. Alpha was tuned by 10-fold cross-validation over five values (0.1, 0.3, 0.5, 0.7, and 0.9). The lambda yielding the minimum cross-validated root mean squared error (RMSE) was selected. Model performance was evaluated in the independent test set using R^2^, RMSE, and Spearman correlation (Appendix 1, Table S7). We further evaluated the stability of the feature selection by performing 1000 bootstrap resampling iterations of the training data. In each bootstrap sample, models were refitted using the tuning parameters selected from the original training set, without additional tuning within the bootstrap loop. The selection frequency of each protein was calculated as the proportion of bootstrap iterations in which each protein was selected. ENET-derived proteomic scores were calculated by multiplying the standardized relative abundance of each aptamer-level protein feature by its corresponding elastic net coefficient, then summing these weighted values for each participant.

For XGB models, limma-based pre-filtering was not applied, as this could restrict the model to features showing linear marginal associations and limit its ability to capture nonlinear patterns and interactions. Therefore, all aptamer-level protein features and covariates were included as predictors. Before modeling, the data were randomly split into training and test sets at an 80:20 ratio. Tuning was conducted in two phases: an exploratory grid search over a broad hyperparameter space and fine-tuning around the best-performing values identified in the exploratory stage (Appendix 2, Supplementary methods 2). Hyperparameters were selected on the training set using stratified 5-fold cross-validation with early stopping to minimize RMSE. We used 5-fold cross-validation because XGB performed staged grid searches making repeated tuning computationally intensive. The final XGB model was trained on the full training set using the best-performing hyperparameters selected during the fine-tuning phase and the corresponding number of boosting rounds identified during cross-validation. Model performance was evaluated in the independent test set using R^2^, RMSE, and Spearman correlation (Appendix 1, Table S7).

To compute protein score from XGB models, we first obtained participant-level predictions from the final XGB model. Then, the predictions were decomposed into protein-specific contributions using SHAP values.^35^ SHAP values indicate the contribution of each protein/feature to an individual’s predicted dietary score. To generate the XGB-derived protein score, the protein-specific SHAP contributions across selected aptamer-level protein features were summed.

### Genetic prioritization using Mendelian randomization

We performed two-sample MR to examine potential causal associations between genetically proxied levels of proteins identified by proteomics discovery analyses with overall cancer, site-specific cancer, T2D, and CVD. Genetic instruments for protein levels were selected from the deCODE genetics plasma proteome resource, which performed a genome-wide association study (GWAS) of plasma protein levels using 4,607 aptamers in 35,556 Icelanders.^36^ We restricted instruments to cis-pQTLs meeting the deCODE study-wide significance threshold of p < 1.8 x 10^-6^.^36^ For each protein, one cis-pQTL instrument was selected, prioritizing variants with complete allele annotation, sentinel conditional signals, absence of high-linkage disequilibrium protein-altering variants, and strong pQTL associations in deCODE.^36^

Outcome GWAS summary statistics for European ancestry were obtained for cancer (overall all cancer),^37^ T2D^38^ and CVD.^36^ We also included site-specific cancers, namely the four most commonly diagnosed globally: breast cancer,^40^ prostate cancer,^41^ lung cancer,^42^ and colorectal cancer,^43^ plus obesity-related cancers, specifically liver cancer,^44^ kidney cancer,^45^ endometrial cancer,^46^ gastric cancer,^44^ and multiple myeloma.^47^ Details of each outcome GWAS, including accession number, sample size, number of cases, and controls, are provided in Appendix 1, Table S31.

As most were instrumented by a single-nucleotide polymorphism (SNP), MR was primarily conducted using the Wald ratio method. When more than one independent cis-pQTL was available, the inverse-variance-weighted MR was applied (six proteins with two SNPs and one protein with three SNPs). Because most analyses relied on single instruments, standard multi-instrument MR sensitivity analyses were not applicable. An FDR < 0.05 was considered significant.

### Colocalization analyses

We conducted Bayesian genetic colocalization analyses to assess whether MR-significant proteins and the outcome disease share a causal variant.^48^ We included SNPs within 500 kb of the instrument variable. For each protein-outcome pair, we estimated posterior probabilities for five hypotheses: H0, the variant has no association with either protein or disease outcome; H1, there was an association with protein only; H2, there was an association with outcome only; H3, there was association with protein and outcome with distinct causal variants; and H4, association with both protein and outcome with a shared causal variant. Posterior probabilities were estimated for each hypothesis by using the default priors in the package coloc.^48^ We focused on the posterior probability for H4 (PP.H4), which estimates the probability that the protein and the disease associations share a common causal variant. PP.H4 ≥ 0.80 suggests strong evidence of colocalization, 0.5 ≤ PP.H4 < 0.80 suggests moderate evidence of colocalization, whereas a PP.H4 < 0.50 suggests weak or no colocalization.

### Further assessment of diet-protein associations in the case-cohort sample

To further assess diet-protein associations among proteins with moderate-to-strong colocalization in a full EPIC proteomics case-cohort sample, we re-examined their associations with the corresponding dietary index through which they were identified, allowing for nonlinearity and effect modification by sex, age, smoking, alcohol consumption, and physical activity. Firstly, we assessed whether age was linearly associated with abundance of each prioritized protein by comparing models with a linear age term against models with restricted cubic splines (RCS) and two to four degrees of freedom. When evidence of age non-linearity was present after FDR correction, the spline’s degrees of freedom with the lowest AIC were selected; otherwise, age was retained as a linear term.

Next, we investigated whether the association between hPDI or uPDI (based on which signature the protein was derived from) and protein abundance was linear, using protein abundance as the outcome and standardized hPDI or uPDI as the exposure. For each protein, we compared a linear dietary index term with RCS models using two to four degrees of freedom, adjusting for the selected age specification and other covariates. When evidence of non-linearity was present after FDR correction, the spline degree of freedom with the lowest AIC was selected; otherwise, the dietary index was retained as a linear term.

Using the selected functional forms for age and dietary index, we tested interactions of dietary index with sex, age, smoking status, alcohol consumption, and physical activity. Interaction p-values were from nested model comparisons corrected for FDR. Analyses were conducted separately for each dietary signature and for the 18 prioritized proteins. All models included sampling weights to account for the case-cohort sampling design.

### Prospective associations of dietary indices, proteomic scores with cancer, T2D, and CVD

We evaluated the associations of protein signatures with three major chronic diseases: overall cancer, T2D, and CVD. Incident overall cancer included cancers across all anatomical sites and was ascertained using ICD-10 codes C00 – C97, excluding non-melanoma skin cancer (C44). Incident T2D was ascertained using code E11, and CVD was ascertained using codes I20 – I25 and I60 – I69. Associations with each incident disease were modeled using accelerated failure time models, with weights to account for oversampling of cases under the case-cohort design, and age as the underlying time scale. Cox proportional hazards models were not used as the proportional hazards assumption was not consistently met, with several proteomic scores and covariates showing evidence of non-proportionality. The event-time distribution was selected separately for each outcome using the Akaike Information Criterion and visual inspection of model fit by comparing fitted survival curves with non-parametric Kaplan–Meier curves. Based on these assessments, Weibull models were selected for overall cancer, whereas generalized gamma models for T2D and CVD.

We first modeled the association between hPDI, uPDI, and each protein score with disease outcome separately. We then fitted mutually adjusted models that included hPDI and each hPDI-related protein score, or uPDI and each uPDI-related protein score, in the same model. All models were adjusted for sex, smoking status, alcohol consumption, physical activity, education, and country of recruitment center.

### Pathway and functional enrichment analyses

We performed functional enrichment analyses for proteins identified by each proteomic discovery approach using Gene Ontology biological processes (GO-BP), cellular components (GO-CC), and molecular function terms (GO-MF), Kyoto Encyclopedia of Genes and Genomes (KEGG), and Reactome. The background universe comprised all aptamer-level protein features available for proteomic discovery. Enriched pathways and functional terms were considered statistically significant at FDR < 0.05.

## Acknowledgements

This work was supported by the National Research Foundation of Korea (NRF) grant funded by the Korea government (Ministry of Science and ICT; grant number RS-2025–00513735 to Jihye Kim).

This study used data from the European Prospective Investigation into Cancer and Nutrition (EPIC) cohort. Coordination of EPIC-Europe is financially supported by International Agency for Research on Cancer (IARC) and Department of Epidemiology and Biostatistics, School of Public Health, Imperial College London with additional infrastructure support from NIHR Imperial Biomedical Research Centre (BRC). The national cohorts are supported by Associazione Italiana per la Ricerca sul Cancro-AIRC-Italy, the Italian Ministry of Health, the Italian Ministry of University and Research (MUR), and Compagnia di San Paolo (Italy); the Dutch Ministry of Public Health, Welfare and Sports (VWS), the Netherlands Organisation for Health Research and Development (ZonMW), and the World Cancer Research Fund (WCRF, The Netherlands); Instituto de Salud Carlos III (ISCIII), the Regional Governments of Andalucía, Asturias, Basque Country, Murcia and Navarra, and the Catalan Institute of Oncology - ICO (Spain); Cancer Research UK (C864/A14136 to EPIC-Norfolk; C8221/A26017 to EPIC-Oxford), Medical Research Council (MR/N003284/1, MC-UU_12015/1 and MC_UU_00006/1 to EPIC-Norfolk; MR/Y013662/1 to EPIC-Oxford) (United Kingdom).

We sincerely acknowledge the Michael J Fox Foundation (#008664 to Christina M. Lill and Elio Riboli), the Cure Alzheimer’s Fund (to Christina M. Lill and Lars Bertram), the ‘CReATe-Clinical Research in ALS and Related Disorders for Therapeutic Development’ Consortium (to Christina M. Lill and Lars Bertram), with additional grant support from the Heisenberg program of the Deutsche Forschungsgemeinschaft (DFG; LI 2654/4 − 1 to Christina M. Lill) in supporting the generation of proteomic data.

SomaScan® data were generated under a Master Research Agreement, 14th December 2021, between Imperial College London and SomaLogic Inc. SomaLogic was not involved in analyzing or interpreting the data, or in writing or submitting the manuscript for publication.

Where authors are identified as personnel of the International Agency for Research on Cancer or WHO, the authors alone are responsible for the views expressed in this Article and they do not necessarily represent the decisions, policy, or views of the International Agency for Research on Cancer or WHO.

## Author contributions

JK and HF conceived the study. JK contributed to the funding acquisition of this study. PL, JK, VV, and HF designed the study. PL developed the analytical approach and planned and performed the statistical analyses. PL, KM, MJS, LPN, DW, QG, VV, PF, JK, and HF provided input to the study design and the statistical analysis. PL drafted the first version of the manuscript and led subsequent revisions in collaboration with all coauthors. KM, MJS, LPN, DW, QG, TT, KP, VV, PF, JK, and HF provided input on an initial draft of the manuscript. All authors contributed substantially to the interpretation of the results, critically revised the manuscript, and provided vital intellectual content. All authors contributed to the writing of the manuscript and approved the final manuscript for submission. JK and HF took responsibility for the integrity of the study data and the accuracy and reproducibility of the analysis.

## Data availability

Data access to EPIC data and/or biospecimens is available via applications.

Instructions for data application are available here: http://epic.iarc.fr/access/index.php. Codes for data analysis will be made available upon request.

## References

1. Carey, C. N. et al. The Environmental Sustainability of Plant-Based Dietary Patterns: A Scoping Review. J. Nutr. 153, 857–869 (2023).

2. Rockström, J. et al. The EAT–Lancet Commission on healthy, sustainable, and just food systems. The Lancet 406, 1625–1700 (2025).

3. Gil-Lespinard, M., Iglesias-Vázquez, L. & Jakszyn, P. Healthful and unhealthful plant-based diets and site-specific cancer risk: a systematic review and meta-analysis of observational studies. Eur. J. Nutr. 65, 121 (2026).

4. Murciano, A. et al. Plant-based diets and risk of type 2 diabetes: systematic review and dose–response meta-analysis. Br. J. Nutr. 134, 277–296 (2025).

5. Satija, A. et al. Healthful and Unhealthful Plant-Based Diets and the Risk of Coronary Heart Disease in U.S. Adults. J. Am. Coll. Cardiol. 70, 411–422 (2017).

6. Zhu, K. et al. Proteomic signatures of healthy dietary patterns are associated with lower risks of major chronic diseases and mortality. Nat. Food 6, 47–57 (2024).

7. Córdova, R. et al. Plant-based dietary patterns and age-specific risk of multimorbidity of cancer and cardiometabolic diseases: a prospective analysis. Lancet Healthy Longev. 6, 100742 (2025).

8. Kim, H. & Rebholz, C. M. Insights from omics research on plant-based diets and cardiometabolic health. Trends Endocrinol. Metab. 36, 546–562 (2025).

9. Kim, H. et al. Plant-Based Diets and Cardiovascular Events: A Proteomics Approach to Examine the Underlying Pathways. J. Nutr. 155, 1741–1750 (2025).

10. Tong, T. Y. N. et al. The plasma proteome of plant-based diets: Analyses of 2920 proteins in 49,615 people. Clin. Nutr. 53, 144–154 (2025).

11. Bray, F. et al. Global cancer statistics 2022: GLOBOCAN estimates of incidence and mortality worldwide for 36 cancers in 185 countries. CA. Cancer J. Clin. 74, 229–263 (2024).

12. Lauby-Secretan, B. et al. Body Fatness and Cancer — Viewpoint of the IARC Working Group. N. Engl. J. Med. 375, 794–798 (2016).

13. Seidah, N. G. & Prat, A. The Multifaceted Biology of PCSK9. Endocr. Rev. 43, 558–582 (2022).

14. Sabatine, M. S. et al. Evolocumab and Clinical Outcomes in Patients with Cardiovascular Disease. N. Engl. J. Med. 376, 1713–1722 (2017).

15. Wang, X. et al. EGFR is a master switch between immunosuppressive and immunoactive tumor microenvironment in inflammatory breast cancer. Sci. Adv. 8, eabn7983 (2022).

16. Malmström, E. et al. Human proteome distribution atlas for tissue-specific plasma proteome dynamics. Cell 188, 2810–2822.e16 (2025).

17. Yang, P., Jiang, X. & He, F. Soluble isoforms of EGFR family in breast cancer: Advances and clinical relevance (Review). Oncol. Lett. 31, 1–14 (2026).

18. Wang, J.-L. et al. Causal relationship between 91 circulating inflammatory proteins and gynecological diseases: A two-sample bidirectional Mendelian randomization study. Int. J. Biol. Macromol. 316, 144729 (2025).

19. Wang, S. E. et al. The Effect of Circulating Proteins and Their Role in Mediating Adiposity’s Effect on Endometrial Cancer Risk: Mendelian Randomization and Colocalization Analyses. Cancer Epidemiol. Biomarkers Prev. 34, 1534–1543 (2025).

20. Gacuta, E. et al. Plasma Levels of Matrilysins (MMP-7 and MMP-26) and Stromelysins (MMP-3 and MMP-10) in Diagnosis of Endometrial Cancer Patients. Int. J. Mol. Sci. 26, 3824 (2025).

21. Retta, S. F. & Glading, A. J. Oxidative stress and inflammation in cerebral cavernous malformation disease pathogenesis: Two sides of the same coin. Int. J. Biochem. Cell Biol. 81, 254–270 (2016).

22. Xia, L. et al. Integrative multi-omics analysis identifies key genes and colocalized signals associated with colorectal cancer risk. BMC Cancer 25, 1372 (2025).

23. Hazelwood, E. et al. Multi-tissue expression and splicing data prioritise anatomical subsite- and sex-specific colorectal cancer susceptibility genes. Nat. Commun. 16, 5043 (2025).

24. Tang, X., Qi, C., Zhou, H. & Liu, Y. Critical roles of PTPN family members regulated by non-coding RNAs in tumorigenesis and immunotherapy. Front. Oncol. 12, 972906 (2022).

25. Zhong, Q. et al. A proteome-wide association study reveals novel plasma proteins as potential therapeutic targets for metabolic dysfunction-associated steatotic liver disease. Front. Endocrinol. 16, 1664691 (2025).

26. Li, Z. et al. Transcriptome and plasma proteome analyses identify susceptibility genes and proteins for alcohol-related hepatocellular carcinoma. Hum. Genomics 10.1186/s40246-026-00984-1 (2026) doi:10.1186/s40246-026-00984-1.

27. Riboli, E. et al. European Prospective Investigation into Cancer and Nutrition (EPIC): study populations and data collection. Public Health Nutr. 5, 1113–1124 (2002).

28. Satija, A. et al. Plant-Based Dietary Patterns and Incidence of Type 2 Diabetes in US Men and Women: Results from Three Prospective Cohort Studies. PLOS Med. 13, e1002039 (2016).

29. Kolijn, P. M. et al. Multi-cohort high-dimensional proteomics reveals early risk markers for lymphoid cancer subtypes. Nat. Commun. 16, 9517 (2025).

30. Rohloff, J. C. et al. Nucleic Acid Ligands With Protein-like Side Chains: Modified Aptamers and Their Use as Diagnostic and Therapeutic Agents. Mol. Ther. - Nucleic Acids 3, e201 (2014).

31. Viallon, V. et al. A New Pipeline for the Normalization and Pooling of Metabolomics Data. Metabolites 11, 631 (2021).

32. Ritchie, M. E. et al. limma powers differential expression analyses for RNA-sequencing and microarray studies. Nucleic Acids Res. 43, e47–e47 (2015).

33. Friedman, J., Hastie, T. & Tibshirani, R. Regularization Paths for Generalized Linear Models via Coordinate Descent. J. Stat. Softw. 33, (2010).

34. Chen, T. & Guestrin, C. XGBoost: A Scalable Tree Boosting System. in Proceedings of the 22nd ACM SIGKDD International Conference on Knowledge Discovery and Data Mining 785–794 (ACM, San Francisco California USA, 2016). doi:10.1145/2939672.2939785.

35. Lundberg, S. M. et al. From local explanations to global understanding with explainable AI for trees. *Nat*. Mach. Intell. 2, 56–67 (2020).

36. Ferkingstad, E. et al. Large-scale integration of the plasma proteome with genetics and disease. Nat. Genet. 53, 1712–1721 (2021).

37. Burrows, K. & Haycock, P. Genome-wide Association Study of Cancer Risk in UK Biobank. University of Bristol 10.5523/BRIS.AED0U12W0EDE20OLB0M77P4B9 (2021).

38. Xue, A. et al. Genome-wide association analyses identify 143 risk variants and putative regulatory mechanisms for type 2 diabetes. Nat. Commun. 9, 2941 (2018).

39. Loh, P.-R., Kichaev, G., Gazal, S., Schoech, A. P. & Price, A. L. Mixed-model association for biobank-scale datasets. Nat. Genet. 50, 906–908 (2018).

40. Michailidou, K. et al. Association analysis identifies 65 new breast cancer risk loci. Nature 551, 92–94 (2017).

41. Wang, A. et al. Characterizing prostate cancer risk through multi-ancestry genome-wide discovery of 187 novel risk variants. Nat. Genet. 55, 2065–2074 (2023).

42. SpiroMeta Consortium et al. Large-scale association analysis identifies new lung cancer susceptibility loci and heterogeneity in genetic susceptibility across histological subtypes. Nat. Genet. 49, 1126–1132 (2017).

43. Fernandez-Rozadilla, C. et al. Deciphering colorectal cancer genetics through multi-omic analysis of 100,204 cases and 154,587 controls of European and east Asian ancestries. Nat. Genet. 55, 89–99 (2023).

44. Verma, A. et al. Diversity and scale: Genetic architecture of 2068 traits in the VA Million Veteran Program. Science 385, eadj1182 (2024).

45. Purdue, M. P. et al. Multi-ancestry genome-wide association study of kidney cancer identifies 63 susceptibility regions. Nat. Genet. 56, 809–818 (2024).

46. O’Mara, T. A. et al. Identification of nine new susceptibility loci for endometrial cancer. Nat. Commun. 9, 3166 (2018).

47. Güler, M. & Canzian, F. Clustering of lymphoid neoplasms by cell of origin, somatic mutation and drug usage profiles: a multi-trait genome-wide association study. Blood Cancer J. 15, 147 (2025).

48. Giambartolomei, C. et al. Bayesian Test for Colocalisation between Pairs of Genetic Association Studies Using Summary Statistics. PLoS Genet. 10, e1004383 (2014).

