## Supplemental Figures for "Candidate Proteins Linking Plant-Based Diet Quality to Cancer, Type 2 Diabetes, and Cardiovascular Disease: Integrating Evidence from Proteomic Analysis and Mendelian Randomization in the EPIC Study"

### Appendix 2

### Supplementary Methods

Figure S1: Volcano plot of linear associations between hPDI and plasma aptamers

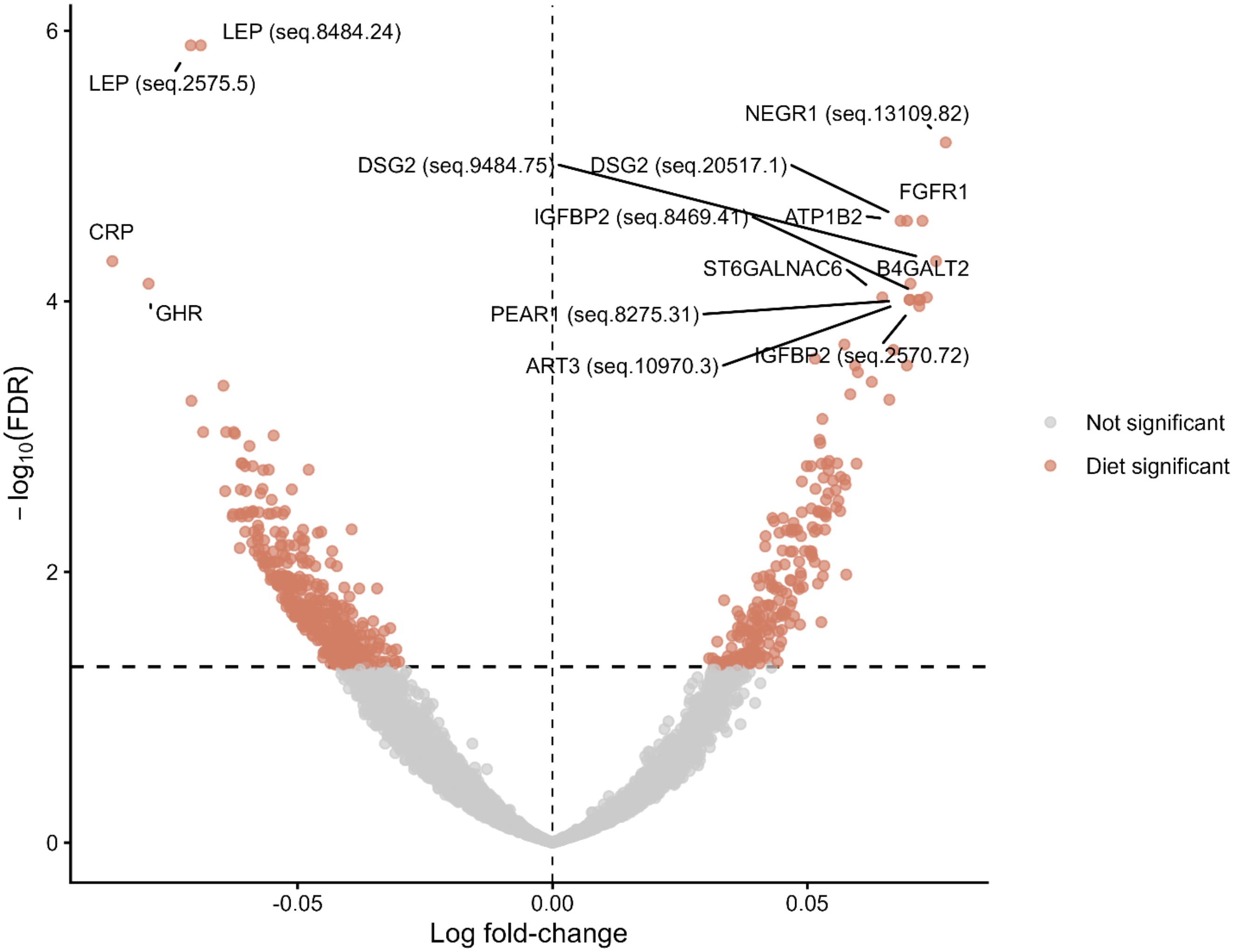

Volcano plot showing linear regression results for the associations between hPDI and plasma aptamer levels. Model was adjusted for age, sex, educational attainment, smoking status, alcohol intake, physical activity, and recruitment center. Each point represents one aptamer. Orange points indicate aptamers associated with hPDI at a false discovery rate-adjusted p-value (FDR) < 0.05, while grey points indicate non-significant aptamers. The dashed horizontal line indicates the FDR threshold of < 0.05. Selected top-ranked aptamers by log fold-change are labelled.

Abbreviation: hPDI – healthful plant-based dietary index, FDR - Benjamini-Hochberg false discovery rate-adjusted p value

Figure S2: Volcano plot of linear associations between uPDI and plasma aptamers

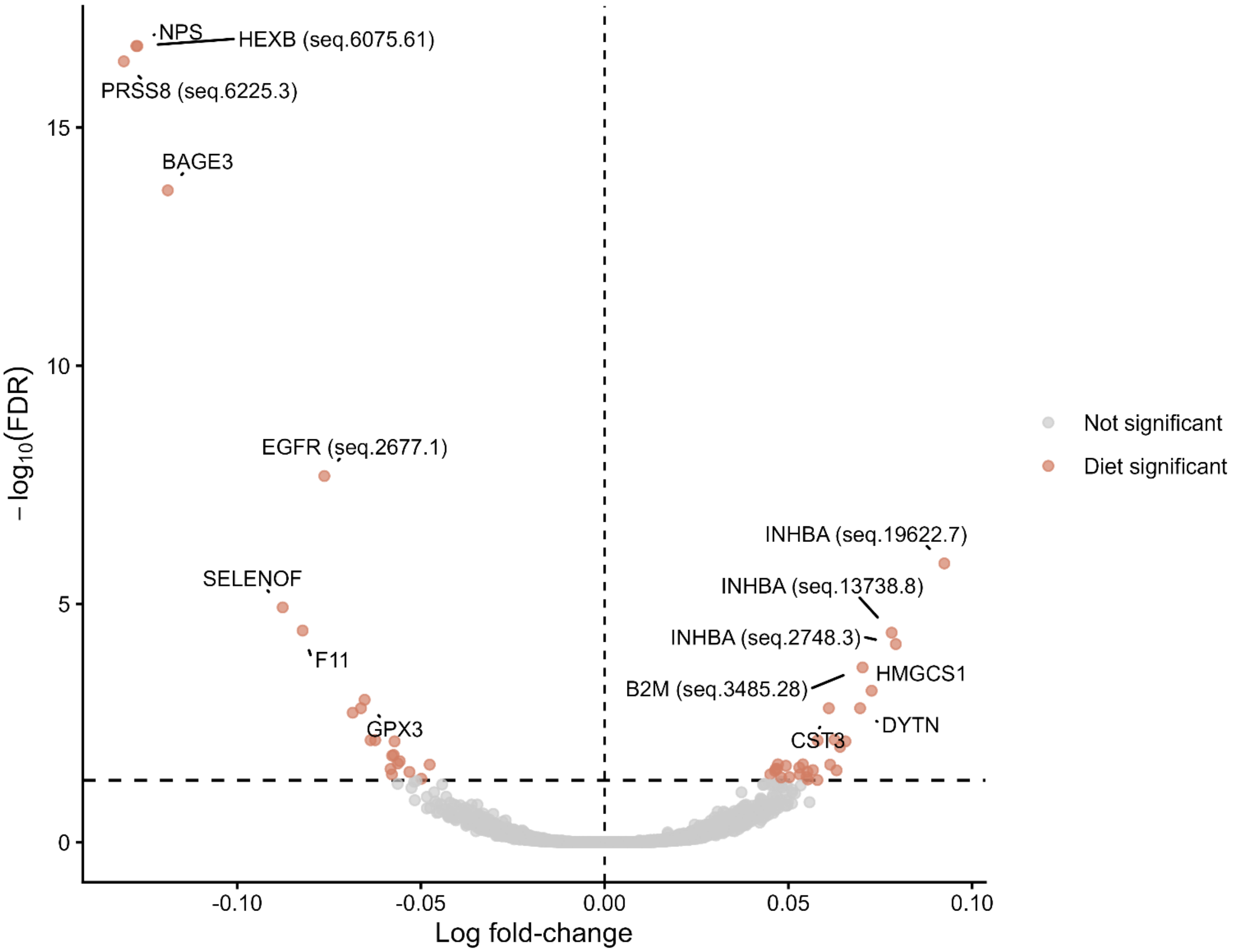

Volcano plot showing linear regression results for the associations between uPDI and plasma aptamer levels. Model was adjusted for age, sex, educational attainment, smoking status, alcohol intake, physical activity, and recruitment center. Each point represents one aptamer. Orange points indicate aptamers associated with uPDI at a false discovery rate-adjusted p-value (FDR) < 0.05, while grey points indicate non-significant aptamers. The dashed horizontal line indicates the FDR threshold of < 0.05. Selected top-ranked aptamers by log fold-change are labelled.

Abbreviation: uPDI – unhealthful plant-based dietary index, FDR - Benjamini-Hochberg false discovery rate-adjusted p value

Figure S3: Top 10 hPDI-related proteins for each signature

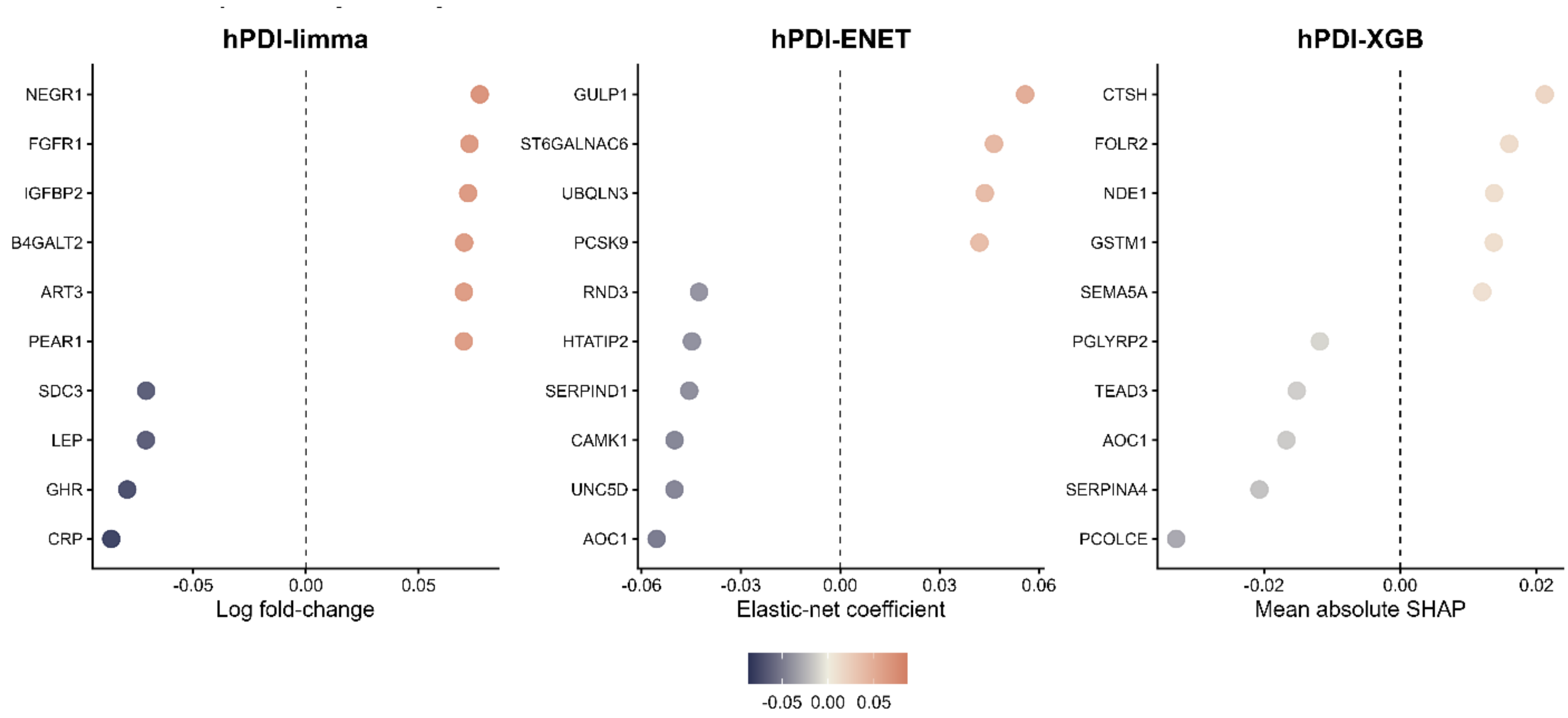

Proteins were ranked by absolute log fold-change for hPDI-limma, absolute coefficient for ENET, and mean absolute SHAP values in the test set for XGB.

Abbreviations: hPDI – healthful plant-based diet index, limma – linear models for microarray, ENET – elastic net, XGB – extreme gradient boosting

Figure S4: Top 10 uPDI-related proteins for each signature

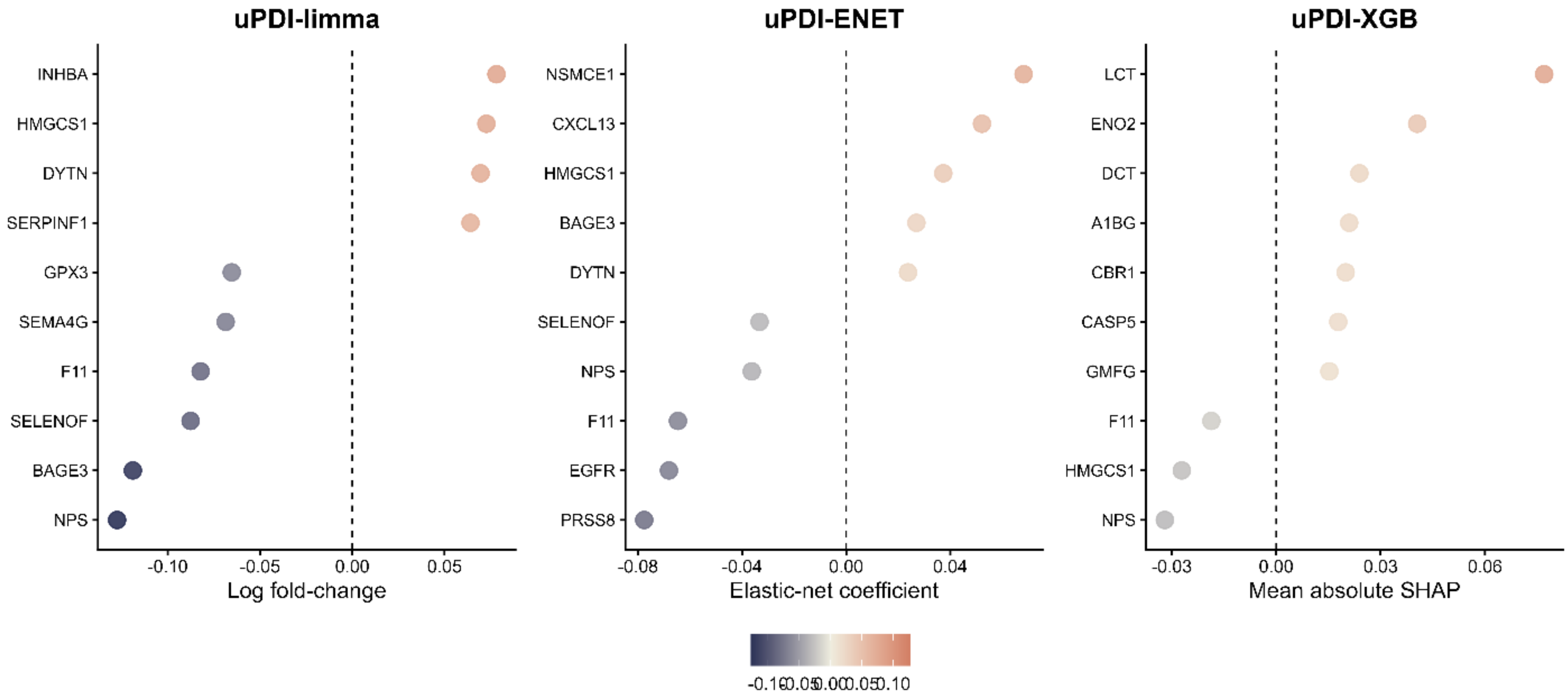

Proteins were ranked by absolute log fold-change for uPDI-limma, absolute coefficient for ENET, and mean absolute SHAP values in the test set for XGB.

Abbreviations: uPDI – unhealthful plant-based diet index, limma – linear models for microarray, ENET – elastic net, XGB – extreme gradient boosting

Figure S5: Protein overlap across hPDI protein signatures

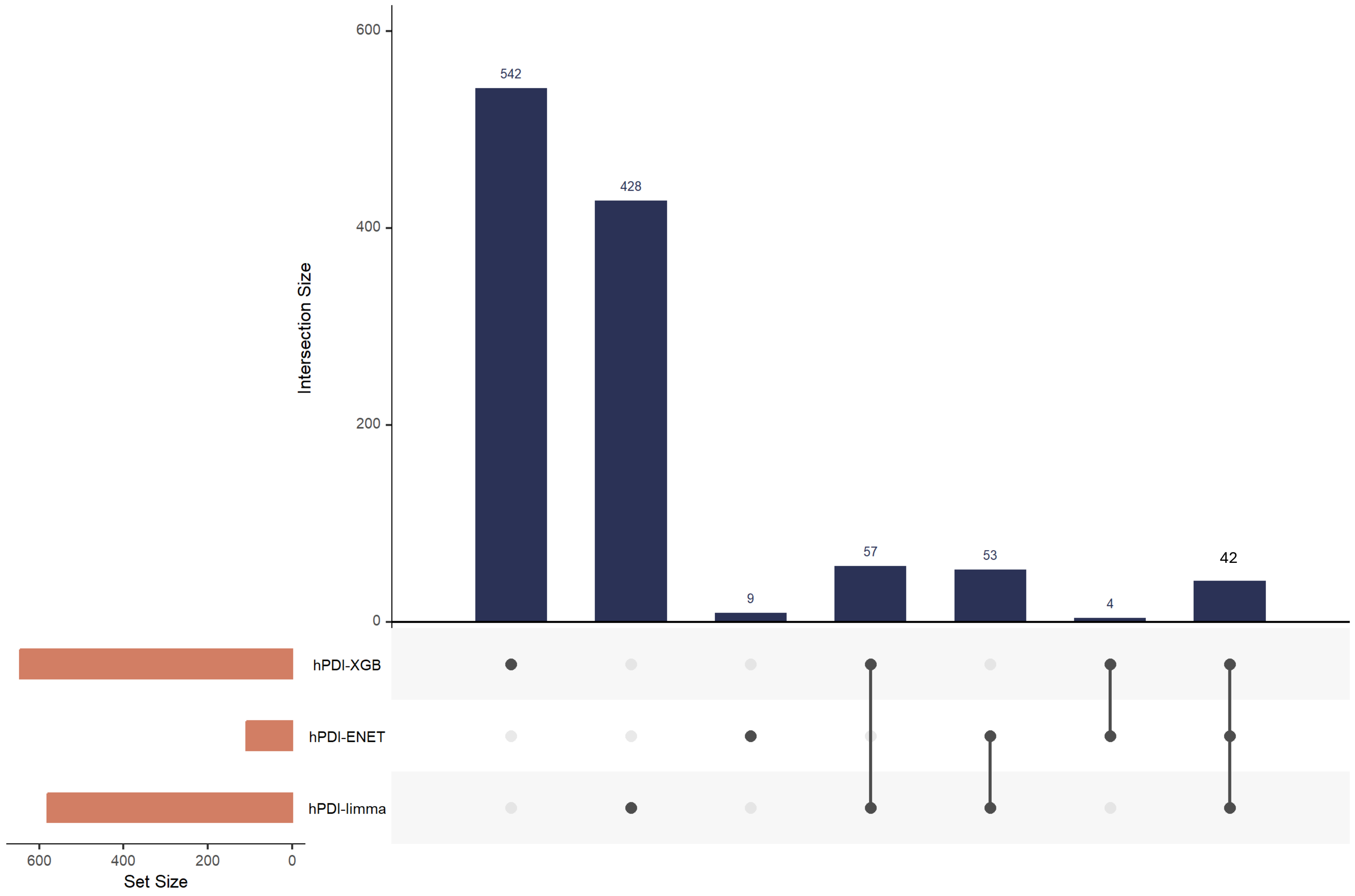

List of aptamers selected by all three signature building approaches:

| Protein | Protein sequence ID | Target | UniProt |
|---|---|---|---|
| PCOLCE | seq.11237.49 | Procollagen C-endopeptidase enhancer 1 | Q15113 |
| EGFLAM | seq.12338.27 | Pikachurin | Q63HQ2 |
| NIPAL4 | seq.12864.9 | Magnesium transporter NIPA4 | Q0D2K0 |
| NEGR1 | seq.13109.82<br>seq.7050.5 | Neuronal growth regulator 1 | Q7Z3B1 |
| SMOC1 | seq.13118.5 | SPARC-related modular calcium-binding protein 1 | Q9H4F8 |
| SEMA5A | seq.13132.14 | Semaphorin-5A | Q13591 |
| GSTM1 | seq.15395.15 | Glutathione S-transferase Mu 1 | P09488 |
| AOC1 | seq.15486.126 | Amiloride-sensitive amine oxidase [copper-containing] | P19801 |
| FOLR2 | seq.15587.20 | Folate receptor beta | P14207 |
| GSN | seq.4775.34<br>seq.16607.78 | Gelsolin | P06396 |
| SDC3 | seq.16612.28 | Syndecan-3 | O75056 |
| APOA4 | seq.17685.9 | Apolipoprotein A-IV | P06727 |
| KIR3DL1 | seq.18907.97 | Killer cell immunoglobulin-like receptor 3DL1 | P43629 |
| RGS21 | seq.19381.7 | Regulator of G-protein signaling 21 | Q2M5E4 |
| VSTM1 | seq.20570.18 | V-set and transmembrane domain-containing protein 1 | Q6UX27 |
| ADGRL3 | seq.20578.10 | Latrophilin-3 | Q9HAR2 |
| DCT | seq.21390.68 | L-dopachrome tautomerase | P40126 |
| UBE2L3\|UBB | seq.21931.27 | UB2L3/PolyUbiquitin K48 | P68036\|<br>P0CG47 |
| IGFBP2 | seq.2570.72<br>seq.22985.160 | Insulin-like growth factor-binding protein 2 | P18065 |
| GHRL | seq.23581.131 | Obestatin | Q9UBU3 |
| MEF2D | seq.24266.2 | Myocyte-specific enhancer factor 2D | Q14814 |
| MAGIX | seq.24696.14 | PDZ domain-containing protein MAGIX | Q9H6Y5 |
| APCS | seq.2474.54 | Serum amyloid P-component | P02743 |

| | | | |
|---|---|---|---|
| PMPCA | seq.24907.3 | Mitochondrial-processing peptidase subunit alpha | Q10713 |
| PPIAL4D | seq.21821.9 | Peptidyl-prolyl cis-trans isomerase A-like 4D | F5H284 |
| IFNL4 | seq.21895.36 | Interferon lambda-4 | K9M1U5 |
| LEP | seq.2575.5 | Leptin | P41159 |
| | seq.8484.24 | | |
| SERPIND1 | seq.3316.58 | Heparin cofactor 2 | P05546 |
| CAMK1 | seq.3592.4 | Calcium/calmodulin-dependent protein kinase type 1 | Q14012 |
| CST5 | seq.3803.10 | Cystatin-D | P28325 |
| NTF4 | seq.4146.58 | Neurotrophin-4 | P34130 |
| C5\|C6 | seq.4482.66 | Complement C5b-C6 complex | P01031\| P13671 |
| UNC5C | seq.5139.32 | Netrin receptor UNC5C | O95185 |
| PCSK9 | seq.5231.79 | Proprotein convertase subtilisin/kexin type 9 | Q8NBP7 |
| | seq.8246.9 | | |
| LEPR | seq.5400.52 | Leptin receptor, soluble | P48357 |
| CRTAC1 | seq.5632.6 | Cartilage acidic protein 1 | Q9NQ79 |
| BPIFA2 | seq.5695.5 | BPI fold-containing family A member 2 | Q96DR5 |
| | seq.16302.11 | | |
| DKK2 | seq.15678.71 | Dickkopf-related protein 2 | Q9UBU2 |
| ST6GALNAC6 | seq.7228.2 | Alpha-N-acetylgalactosaminide alpha-2,6-sialyltransferase 6 | Q969X2 |
| NAALAD2 | seq.7986.98 | N-acetylated-alpha-linked acidic dipeptidase 2 | Q9Y3Q0 |
| PEAR1 | seq.8275.31 | Platelet endothelial aggregation receptor 1:Extracellular domain | Q5VY43 |
| LILRA4 | seq.8299.66 | Leukocyte immunoglobulin-like receptor subfamily A member 4 | P59901 |
| BST2 | seq.8832.55 | Bone marrow stromal antigen 2 | Q10589 |

Abbreviation: hPDI – healthful plant-based diet index, limma – linear models for microarray, ENET – elastic net, XGB – extreme gradient boosting

Figure S6: Protein overlap across uPDI protein signatures

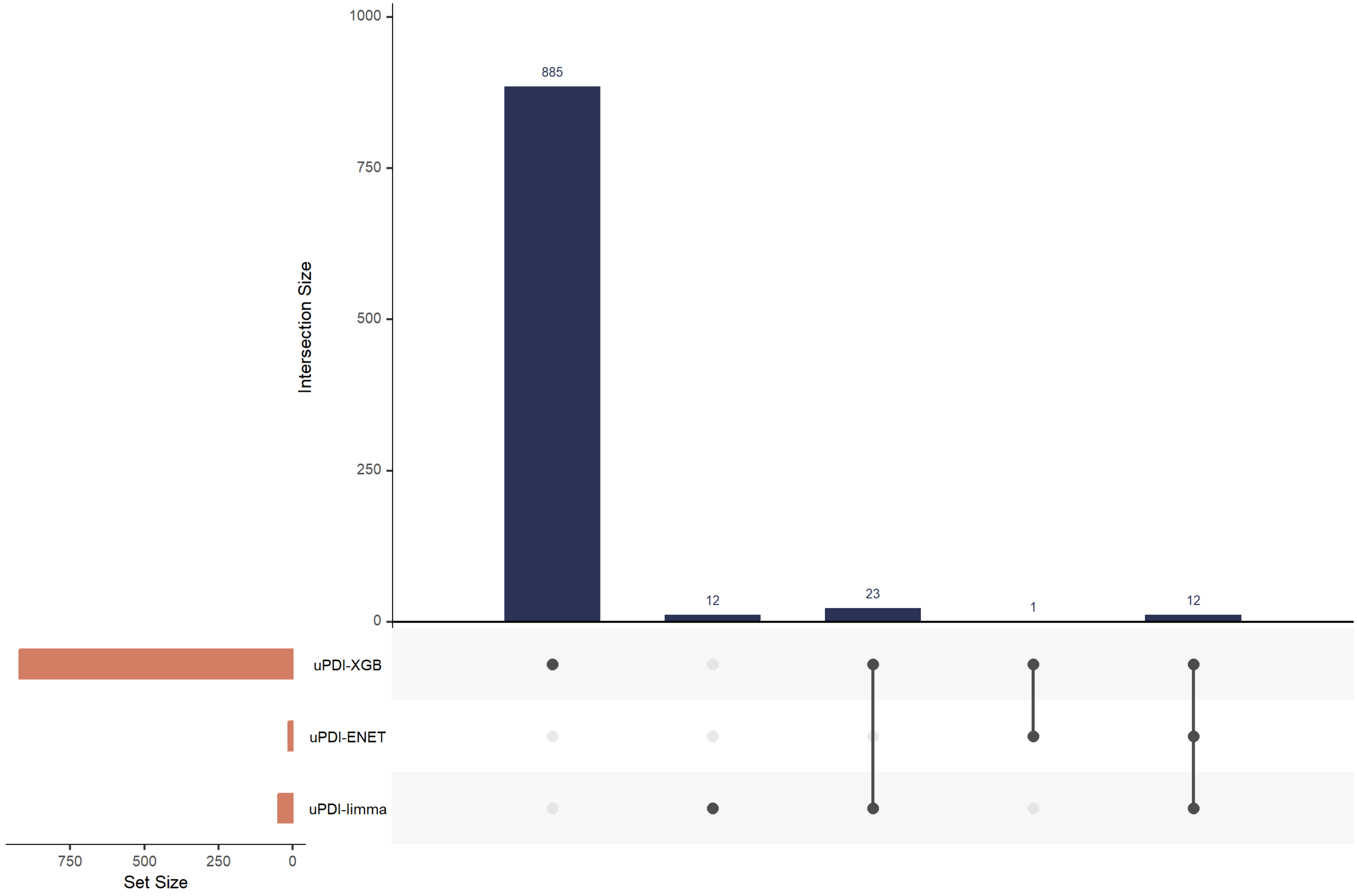

List of aptamers selected by all three signature building approaches:

<table>
<tr><th>Protein</th><th>Protein sequence ID</th><th>Target</th><th>UniProt</th></tr>
<tr><td>HMGCS1</td><td>seq.13496.19</td><td>Hydroxymethylglutaryl-CoA synthase, cytoplasmic</td><td>Q01581</td></tr>
<tr><td rowspan="2">CXCL13</td><td>seq.3487.32</td><td rowspan="2">C-X-C motif chemokine 13</td><td rowspan="2">O43927</td></tr>
<tr><td>seq.13701.2</td></tr>
<tr><td rowspan="3">INHBA</td><td>seq.19622.7</td><td rowspan="2">Activin A</td><td rowspan="3">P08476</td></tr>
<tr><td>seq.2748.3</td></tr>
<tr><td>seq.13738.8</td><td>Inhibin beta A chain</td></tr>
<tr><td>SELENOF</td><td>seq.20087.3</td><td>15 kDa selenoprotein</td><td>O60613</td></tr>
<tr><td>F11</td><td>seq.2190.55</td><td>Coagulation Factor XI</td><td>P03951</td></tr>
<tr><td>DYTN</td><td>seq.22124.94</td><td>Dystrotelin</td><td>A2CJ06</td></tr>
<tr><td>LIMK1</td><td>seq.25308.8</td><td>LIM domain kinase 1</td><td>P53667</td></tr>
<tr><td>EGFR</td><td>seq.2677.1</td><td>Epidermal growth factor receptor</td><td>P00533</td></tr>
<tr><td>PRSS8</td><td>seq.6225.3</td><td>Prostasin</td><td>Q16651</td></tr>
<tr><td>NPS</td><td>seq.6390.18</td><td>Neuropeptide S</td><td>P0C0P6</td></tr>
<tr><td>BAGE3</td><td>seq.6442.6</td><td>B melanoma antigen 3</td><td>Q86Y29</td></tr>
<tr><td>NPW</td><td>seq.9986.14</td><td>Neuropeptide W</td><td>Q8N729</td></tr>
</table>

Abbreviation: uPDI – unhealthful plant-based diet index, limma – linear models for microarray, ENET – elastic net, XGB – extreme gradient boosting

Figure S7: Heatmap of hPDI-related proteins significantly associated with disease outcomes by Mendelian randomization

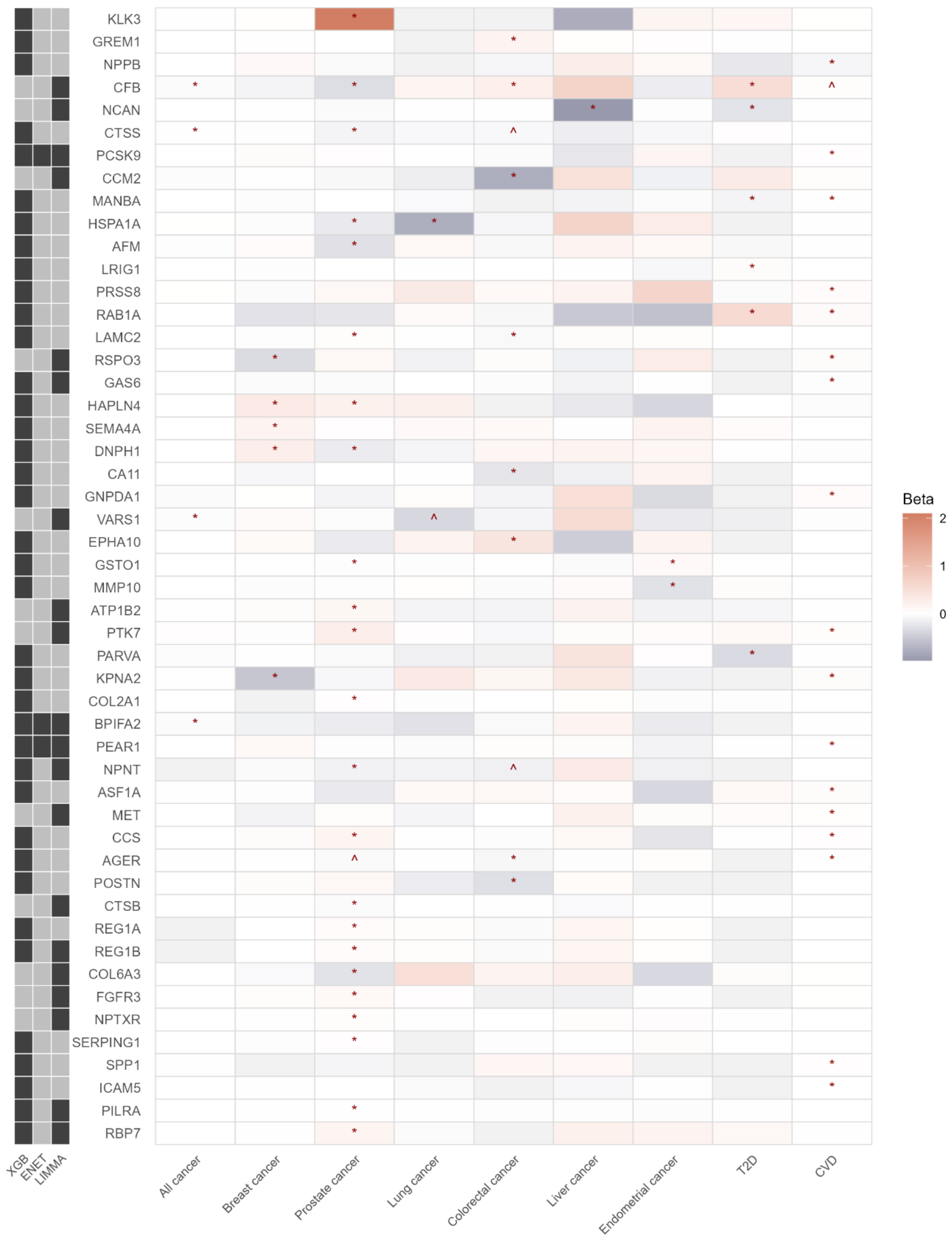

Heatmap of hPDI-related proteins in association with diseases. Rows represent prioritized proteins and columns represent disease outcomes, including overall cancer, site-specific cancers, T2D, and CVD. Cell color indicates beta estimates for the association between each protein and disease outcomes, with blue indicating negative values, red indicating positive values, white indicating null or weaker associations. Symbols denote statistical significance after multiple-testing correction (* - FDR < 0.05, ^ FDR < 0.01). The left annotation panel indicates the method of protein selection by which each protein was identified, with dark/black shading indicating the selection by the corresponding method.

Abbreviations: limma - proteins selected in linear models for microarray data, ENET - proteins selected in elastic net, hPDI-XGB - proteins selected in extreme gradient boosting, FDR – false discovery rate, Benjamini-Hochberg-adjusted p-value

Figure S8: Heatmap of uPDI-related proteins significantly associated with disease outcomes by Mendelian randomization

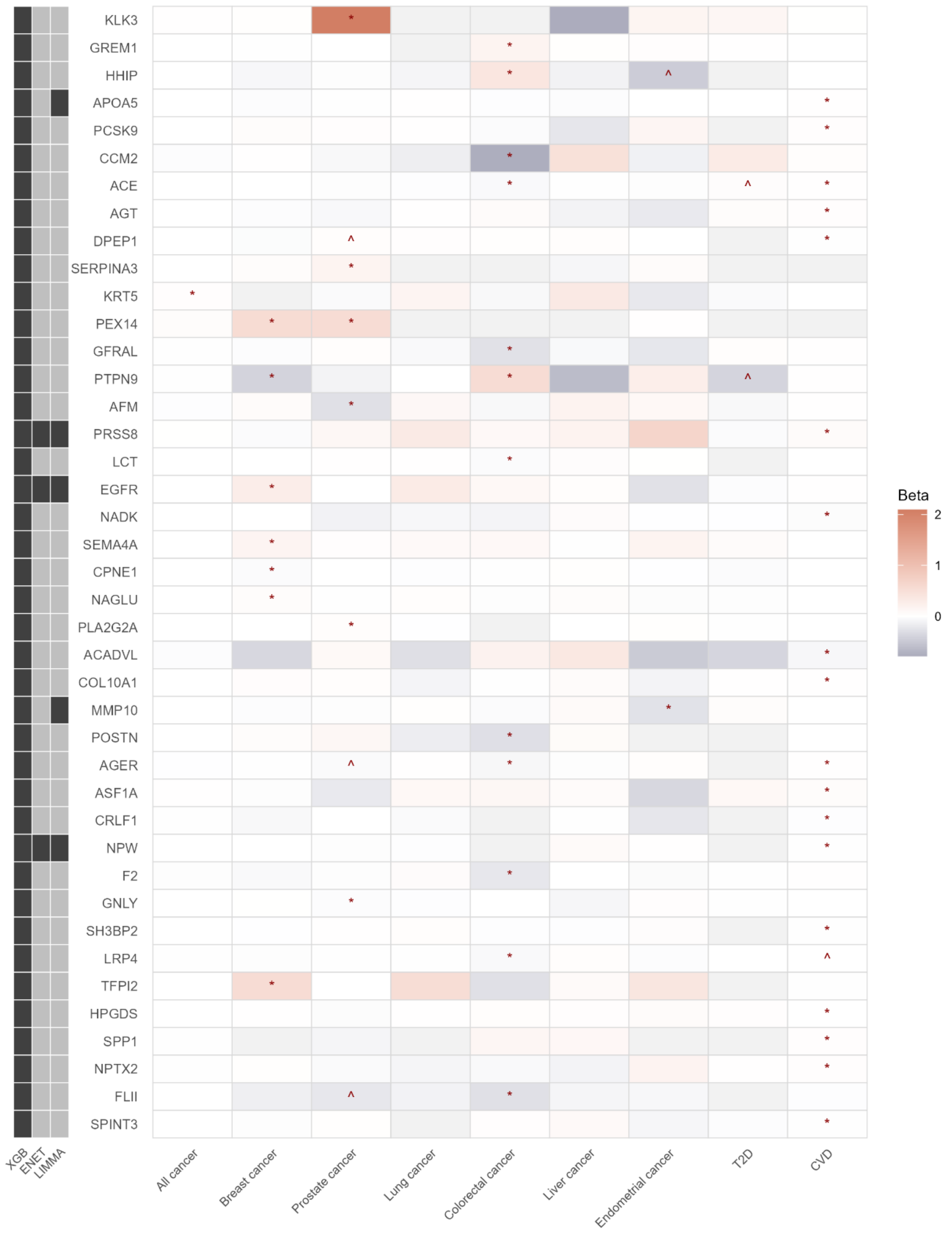

Heatmap of prioritized uPDI-related proteins in association with diseases. Rows represent prioritized proteins and columns represent disease outcomes, including overall cancer, site-specific cancers, T2D, and CVD. Cell color indicates beta estimates for the association between each protein and disease outcomes, with blue indicating negative values, red indicating positive values, white indicating null or weaker associations. Symbols denote statistical significance after multiple-testing correction (* - FDR < 0.05, ^ FDR < 0.01). The left annotation panel indicates the method of protein selection by which each protein was identified, with dark/black shading indicating the selection by the corresponding method.

Abbreviations: limma - proteins selected in linear models for microarray data, ENET - proteins selected in elastic net, uPDI-XGB - proteins selected in extreme gradient boosting, FDR – false discovery rate, Benjamini-Hochberg-adjusted p-value, T2D – type 2 diabetes, CVD – cardiovascular disease

Figure S9: Associations between hPDI and abundance of hPDI-related proteins with colocalized evidence by sex

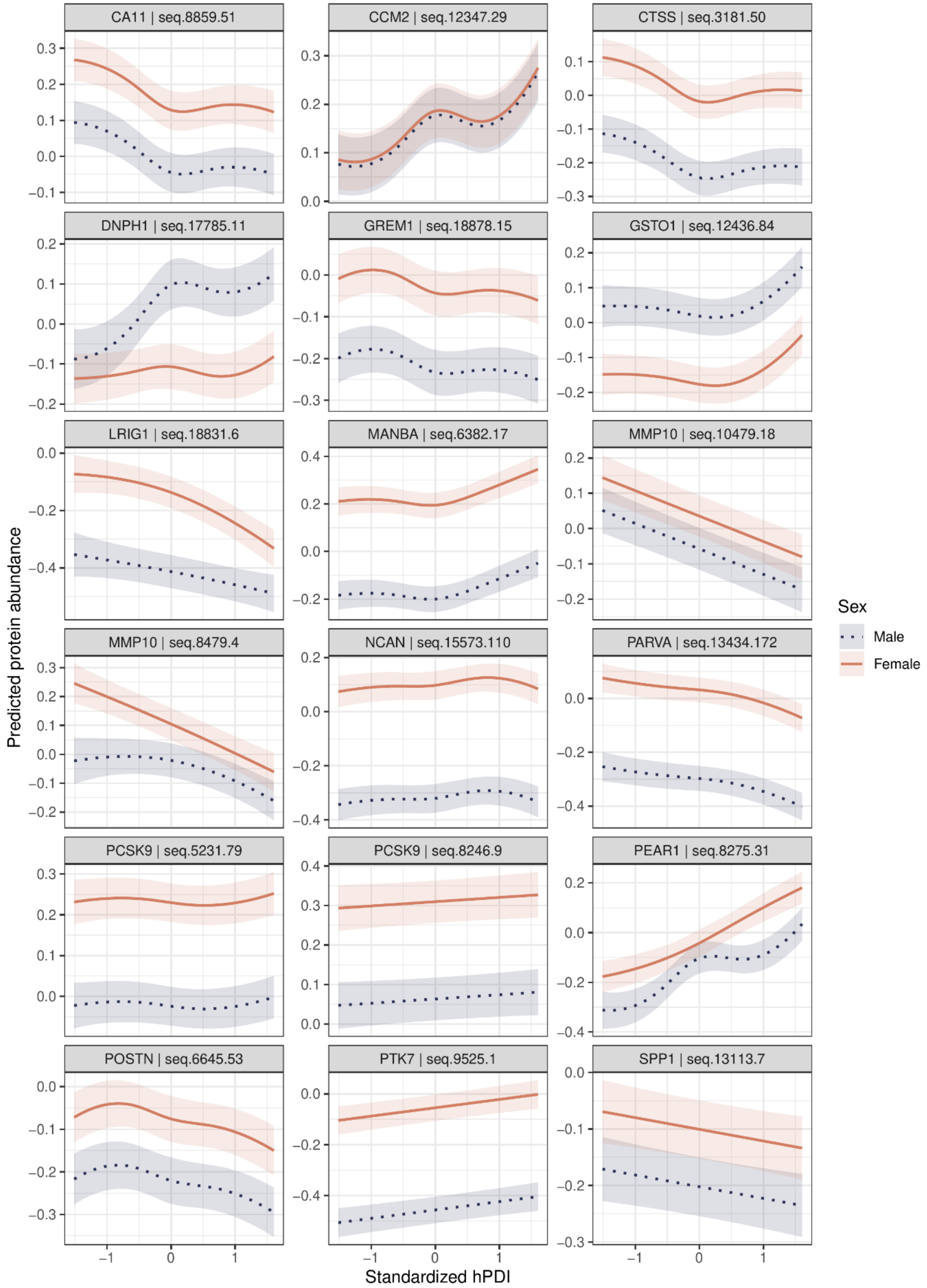

Panels show predicted protein abundance across standardized hPDI values, stratified by sex. Models were adjusted for age, educational attainment, smoking status, alcohol intake, physical activity, and country of recruitment. Age was specified as restricted cubic splines when evidence of nonlinearity was present. The dotted blue line represents males, and the solid orange line represents females. Shaded bands indicate 95% confidence intervals. Each panel represents one protein, and y-axes are scaled separately.

Abbreviations: hPDI – healthful plant-based diet index

Figure S10: Associations between uPDI and abundance of uPDI-related proteins with colocalized evidence by sex

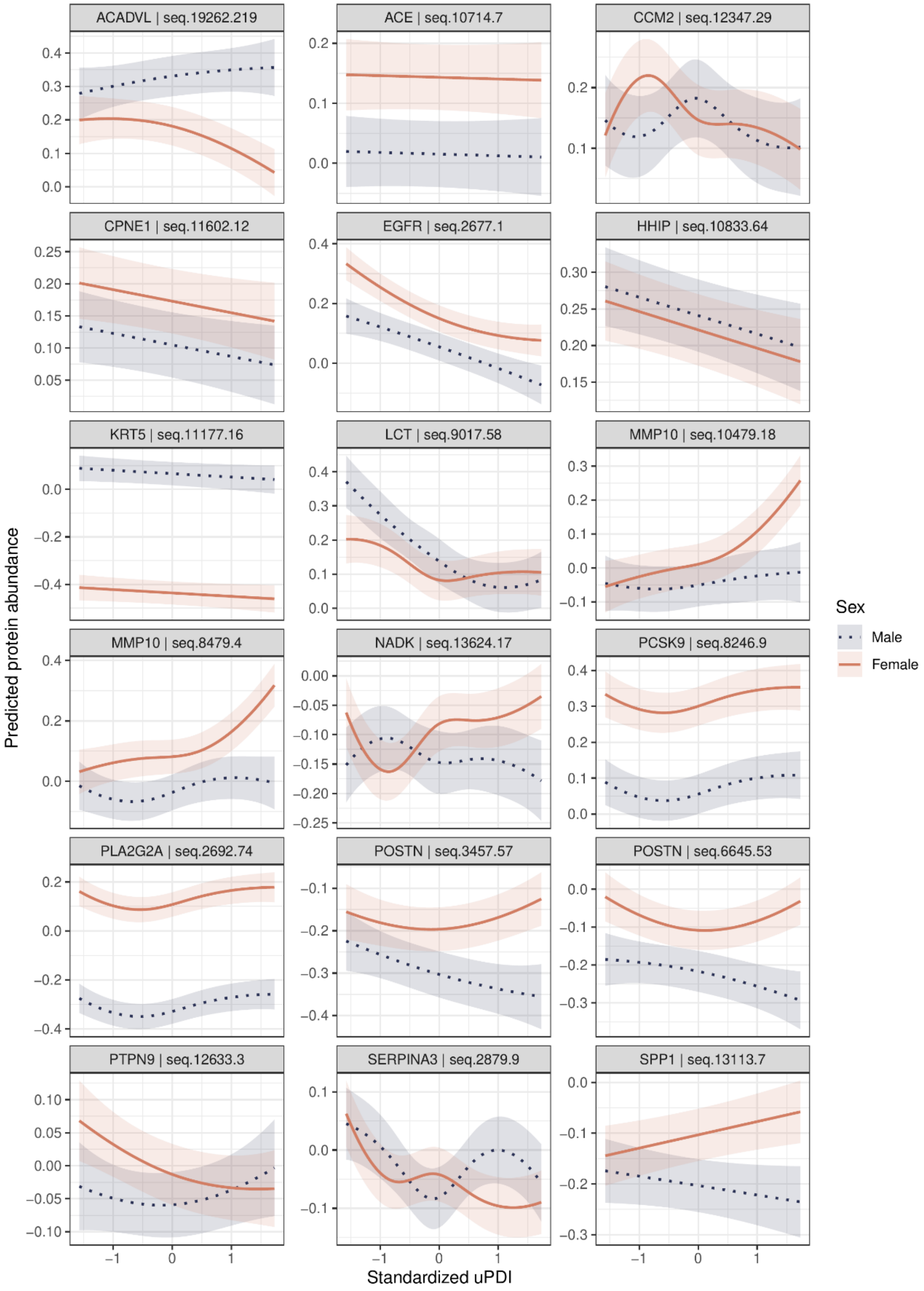

Panels show predicted protein abundance across standardized uPDI values, stratified by sex. Models were adjusted for age, educational attainment, smoking status, alcohol intake, physical activity, and country of recruitment. Age was specified as a restricted cubic spline when evidence of nonlinearity was present. The dotted blue line represents males, and the solid orange line represents females. Shaded bands indicate 95% confidence intervals. Each panel represents one protein, and y-axes are scaled separately.

Abbreviations: uPDI – unhealthful plant-based diet index

Figure S11: Associations between hPDI and abundance of hPDI-related proteins with colocalized evidence by smoking status

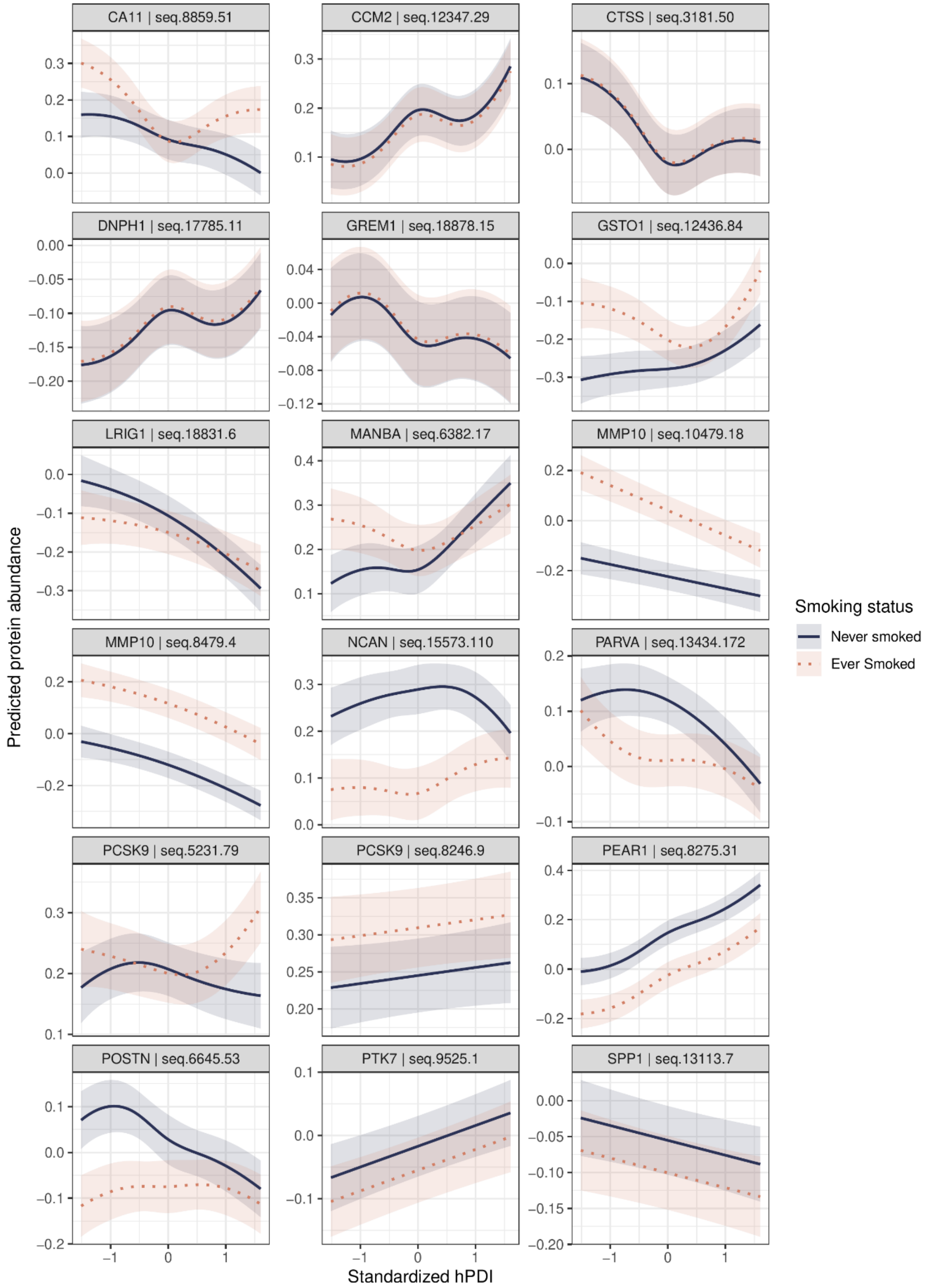

Panels show predicted protein abundance across standardized hPDI values, stratified by smoking status. Models were adjusted for age, educational attainment, sex, alcohol intake, physical activity, and country of recruitment. Age was specified as a restricted cubic spline when evidence of nonlinearity was present. The solid blue line represents never smokers, and the dotted orange lines represent ever smokers. Shaded bands indicate 95% confidence intervals. Each panel represents one protein, and y-axes are scaled separately.

Abbreviations: hPDI – healthful plant-based diet index

Figure S12: Associations between hPDI and abundance of hPDI-related proteins with colocalized evidence by alcohol consumption

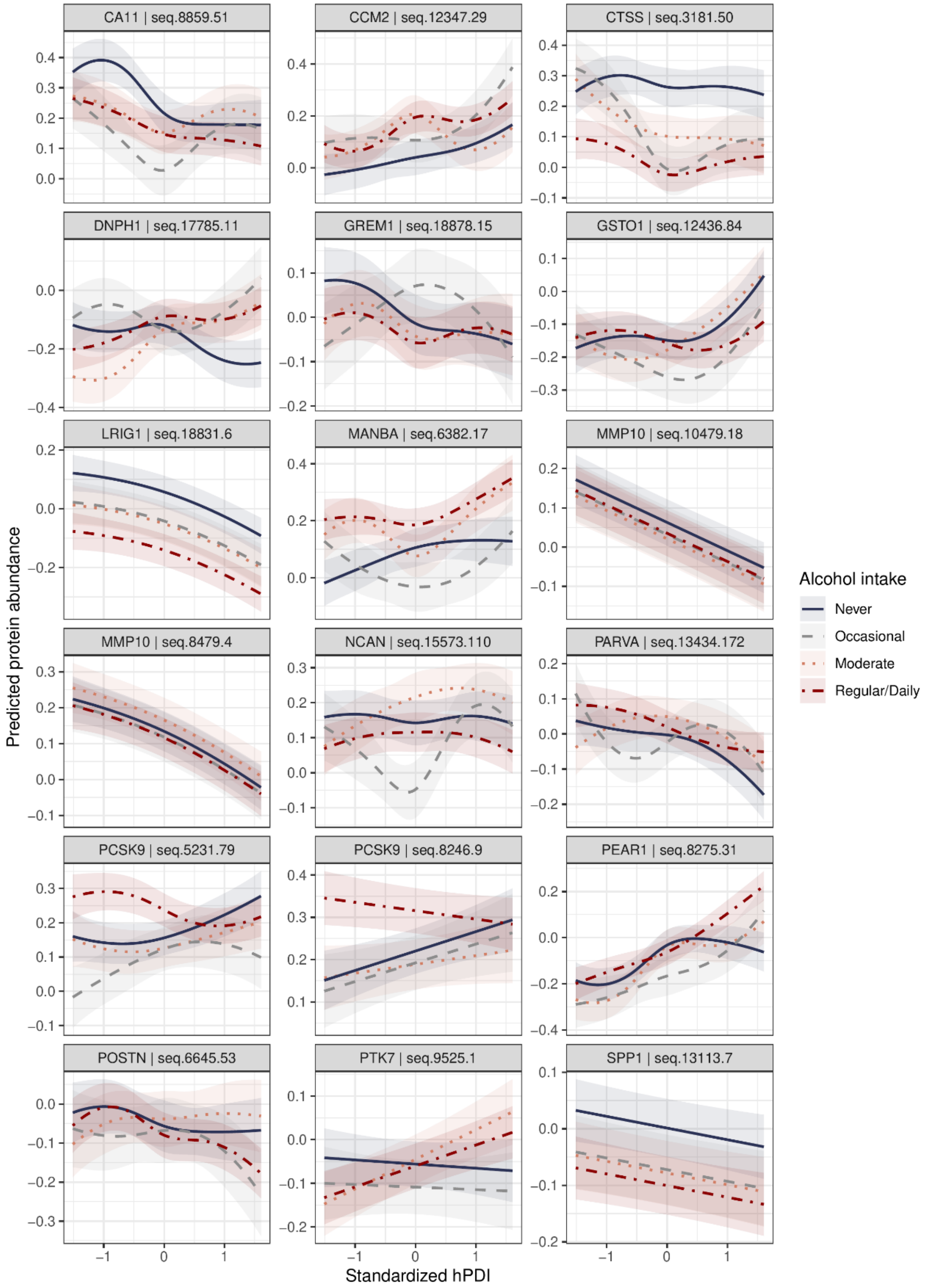

Panels show predicted protein abundance across standardized hPDI values, stratified by alcohol consumption. Models were adjusted for age, educational attainment, sex, smoking status, physical activity, and country of recruitment. Age was specified as a restricted cubic spline when evidence of nonlinearity was present. The solid blue line represents those who never drank, the dashed gray line represents occasional drinkers, the orange dotted line represents moderate drinkers, and the dashed red line represents regular or daily drinkers. Shaded bands indicate 95% confidence intervals. Each panel represents one protein, and y-axes are scaled separately.

Abbreviations: hPDI – healthful plant-based diet index

Figure S13: Associations between hPDI and abundance of hPDI-related proteins with colocalized evidence by age

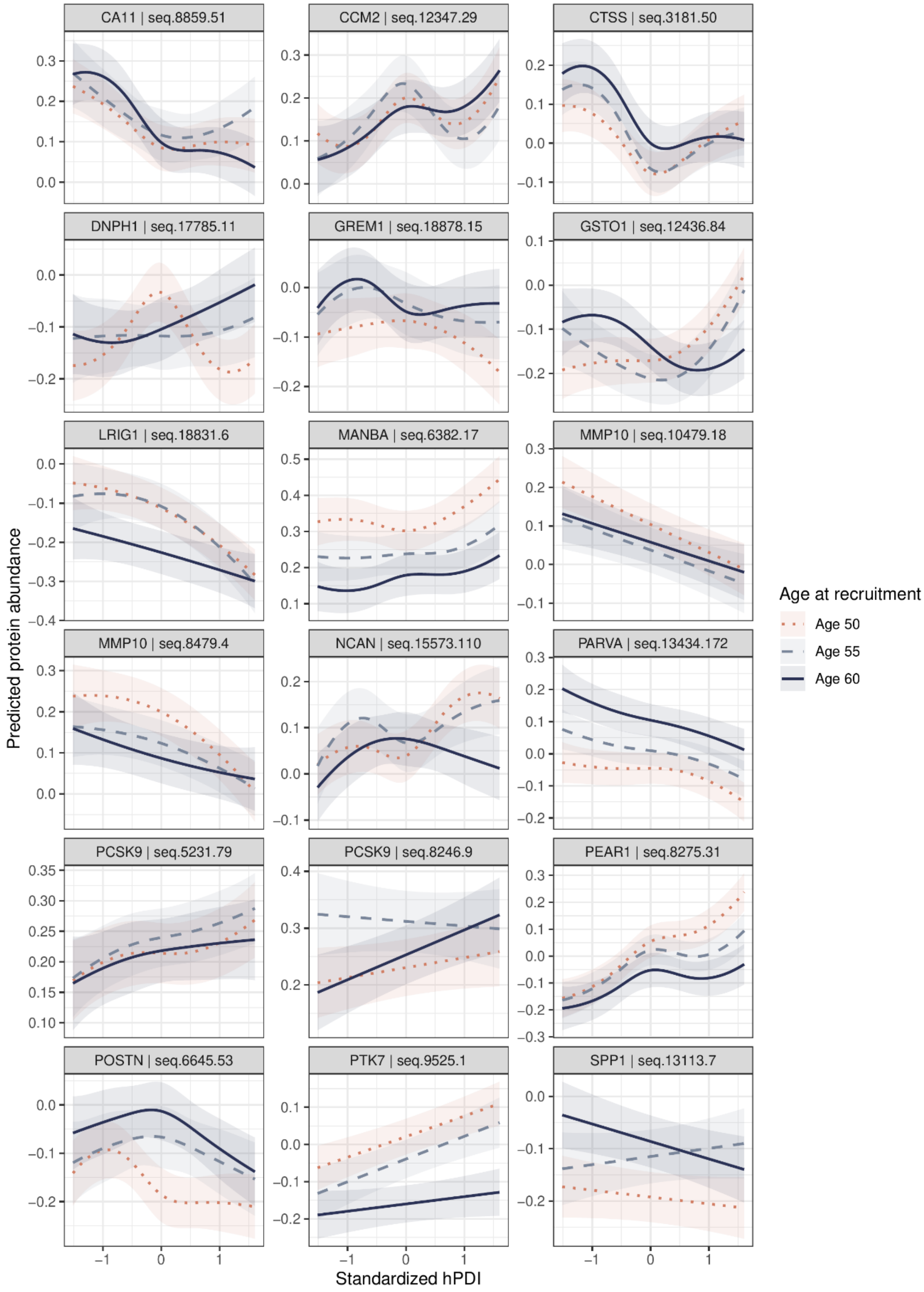

Panels show predicted protein abundance across standardized hPDI values for ages 50, 55, and 60 years. Models were adjusted for educational attainment, sex, smoking status, alcohol consumption, physical activity, and country of recruitment. Age was specified as a restricted cubic spline when evidence of nonlinearity was present. The orange dotted lines represent predictions made at age 50, the dashed gray lines represent predictions at age 55, and the solid blue lines represent predictions at age 60. Shaded bands indicate 95% confidence intervals. Each panel represents one protein, and y-axes are scaled separately.

Abbreviations: hPDI – healthful plant-based diet index

Figure S14: Associations between hPDI and abundance of hPDI-related proteins with colocalized evidence by physical activity levels

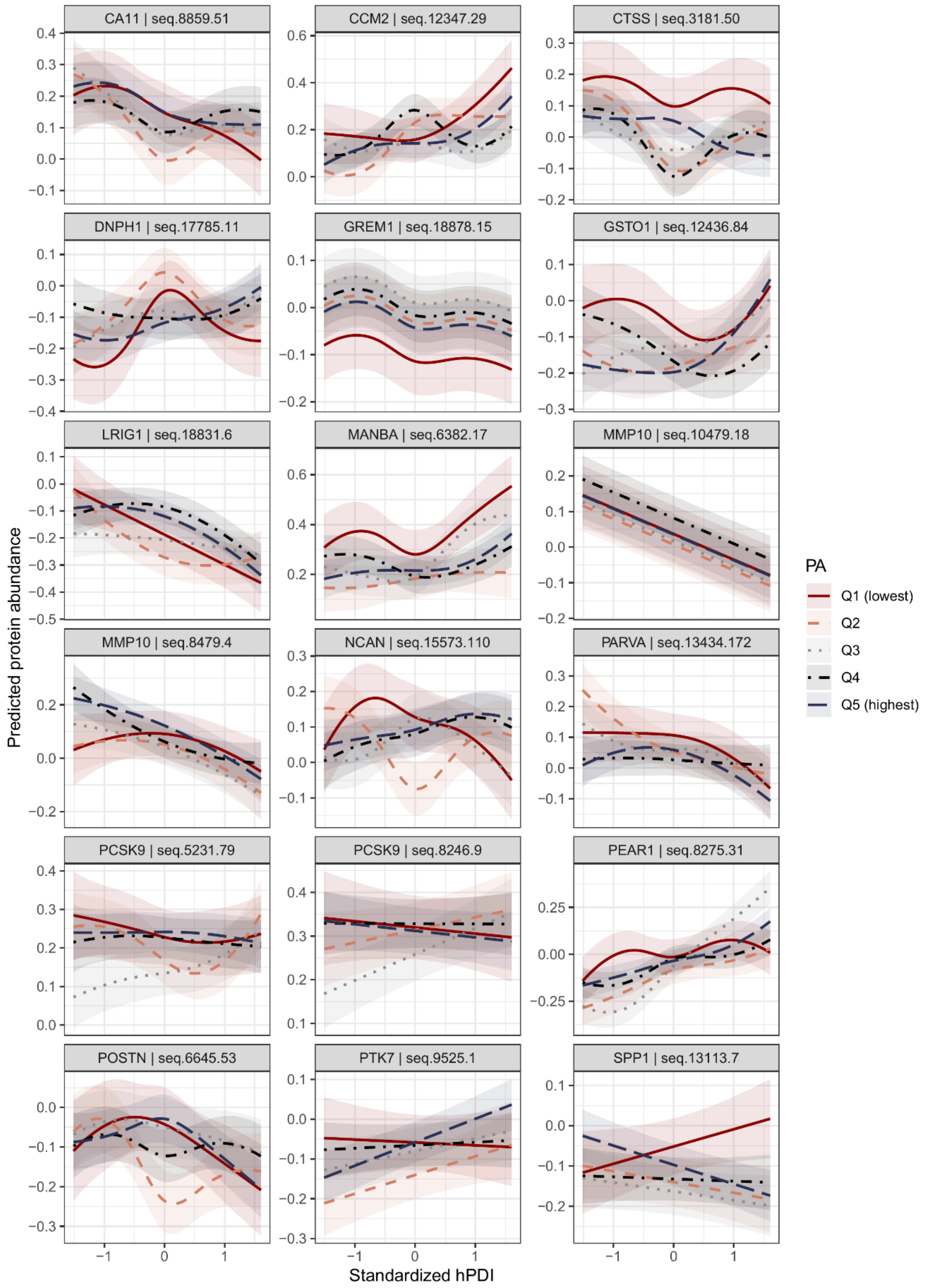

Panels show predicted protein abundance across standardized hPDI values, stratified by quintiles of physical activity levels. Models were adjusted for age, educational attainment, sex, smoking status, alcohol consumption, and country of recruitment. Age was specified as restricted cubic splines when evidence of nonlinearity was present. Lines represent physical activity quintiles (Q1–Q5), as indicated in the legend. Shaded bands indicate 95% confidence intervals. Each panel represents one protein, and y-axes are scaled separately.

Abbreviations: hPDI – healthful plant-based diet index, PA – physical activity, Q1- first quintile, lowest level of physical activity, Q2 – second quintile, Q3 – third quintile, Q4 - fourth quintile, Q5 - fifth quintile, highest level of physical activity

Figure S15: Associations between uPDI and abundance of uPDI-related proteins with colocalized evidence by smoking status

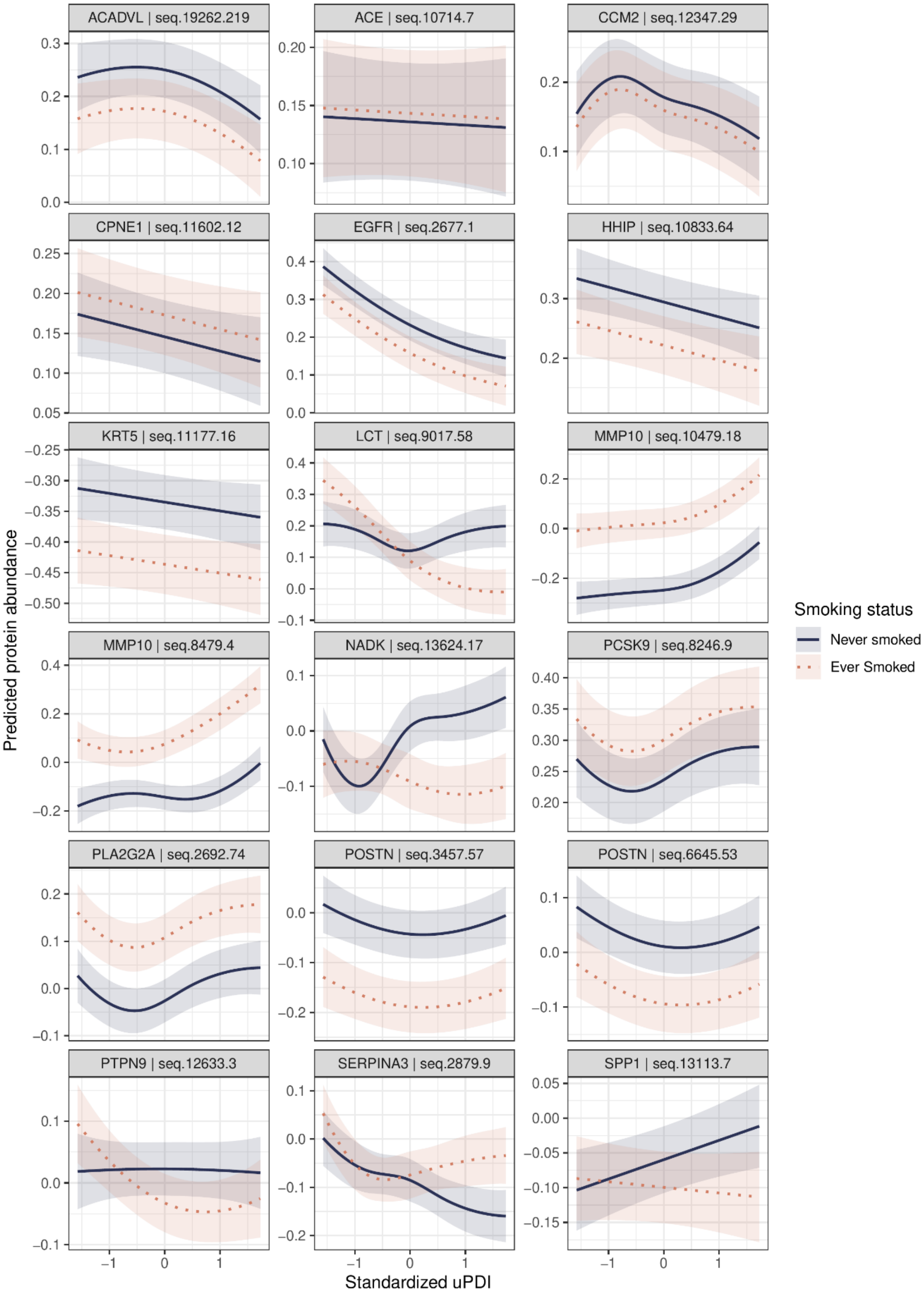

Panels show predicted protein abundance across standardized uPDI values, stratified by smoking status. Models were adjusted for age, educational attainment, sex, alcohol intake, physical activity, and country of recruitment. Age was specified as a restricted cubic spline when evidence of nonlinearity was present. The solid blue line represents never smokers, and the dotted orange lines represent ever smokers. Shaded bands indicate 95% confidence intervals. Each panel represents one protein, and y-axes are scaled separately.

Abbreviations: uPDI – unhealthful plant-based diet index

Figure S16: Associations between uPDI and abundance of uPDI-related proteins with colocalized evidence by alcohol consumption

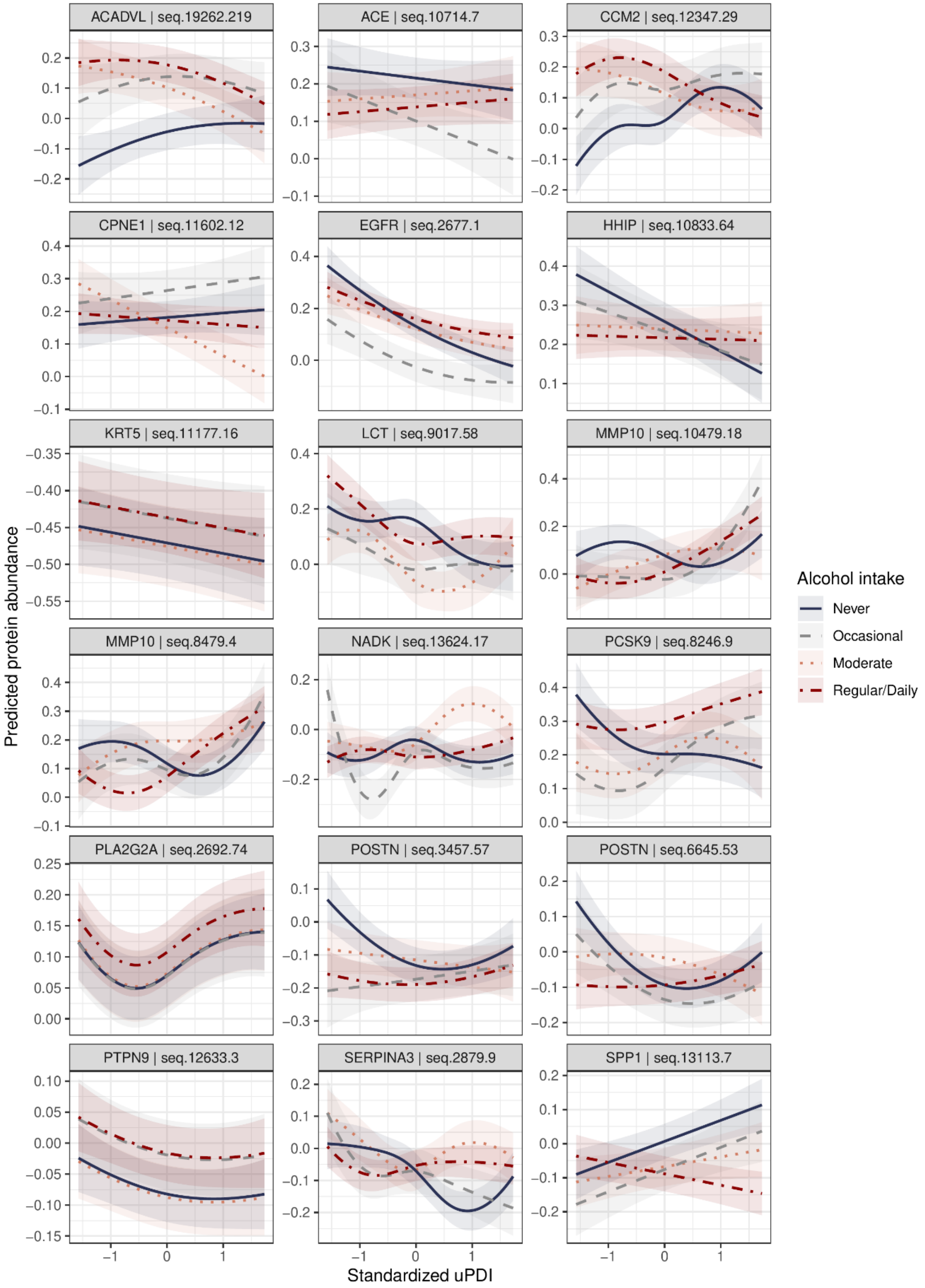

Panels show predicted protein abundance across standardized uPDI values, stratified by alcohol consumption. Models were adjusted for age, educational attainment, sex, smoking status, physical activity, and country of recruitment. Age was specified as restricted cubic splines when evidence of nonlinearity was present. The solid blue line represents those who never drank, the dashed gray line represents occasional drinkers, the orange dotted line represents moderate drinkers, and the dashed red line represents regular or daily drinkers. Shaded bands indicate 95% confidence intervals. Each panel represents one protein, and y-axes are scaled separately.

Abbreviations: uPDI – unhealthful plant-based diet index

Figure S17: Associations between uPDI and abundance of uPDI-related proteins with colocalized evidence by age

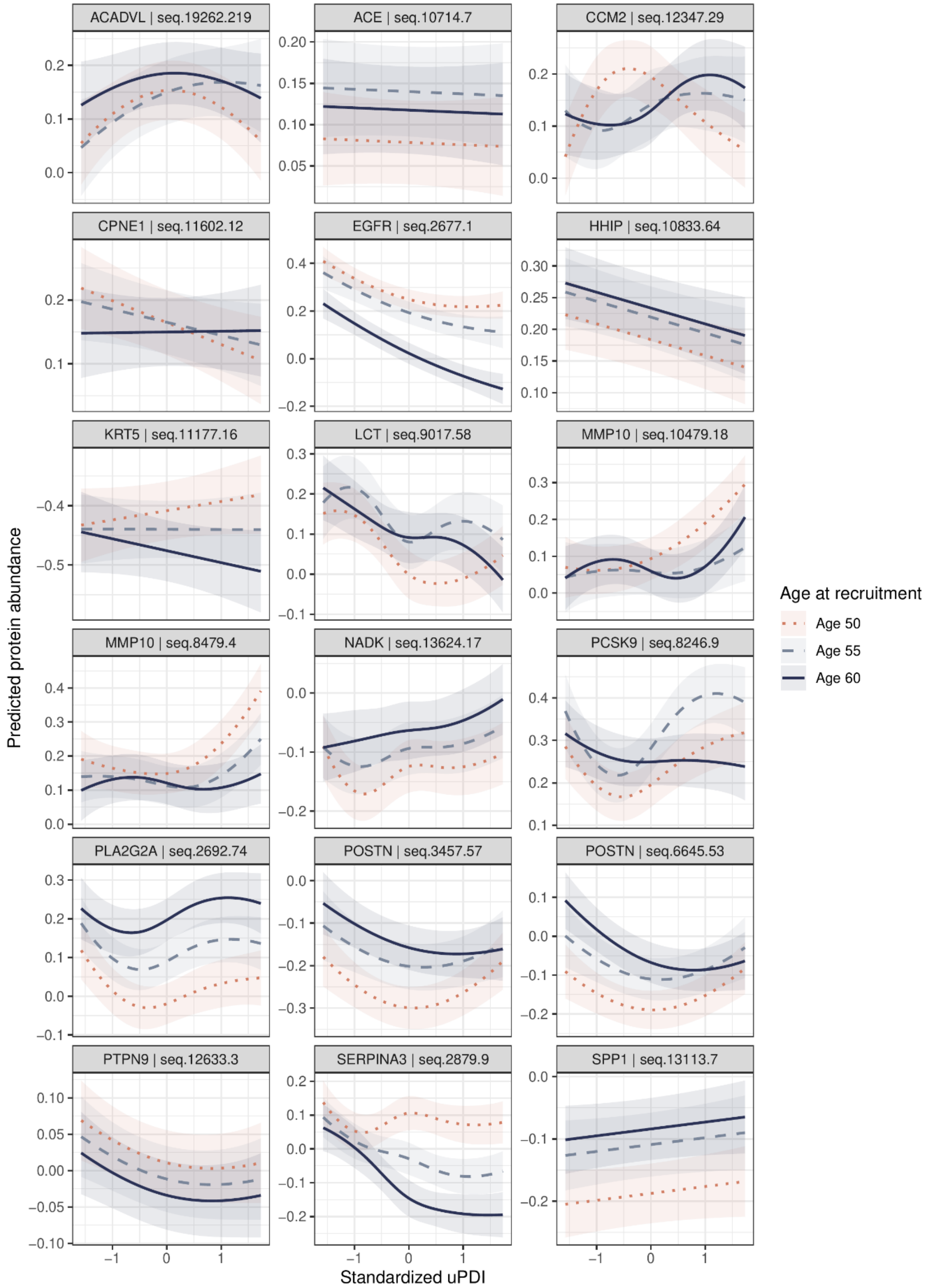

Panels show predicted protein abundance across standardized uPDI values for ages 50, 55, and 60 years. Models were adjusted for educational attainment, sex, smoking status, alcohol consumption, physical activity, and country of recruitment. Age was specified as a restricted cubic spline when evidence of nonlinearity was present. The orange dotted lines represent predictions made at age 50, the dashed gray lines represent predictions at age 55, and the solid blue lines represent predictions at age 60. Shaded bands indicate 95% confidence intervals. Each panel represents one protein, and y-axes are scaled separately.

Abbreviations: uPDI – unhealthful plant-based diet index

Figure S18: Associations between uPDI and abundance of uPDI-related proteins with colocalized evidence by physical activity levels

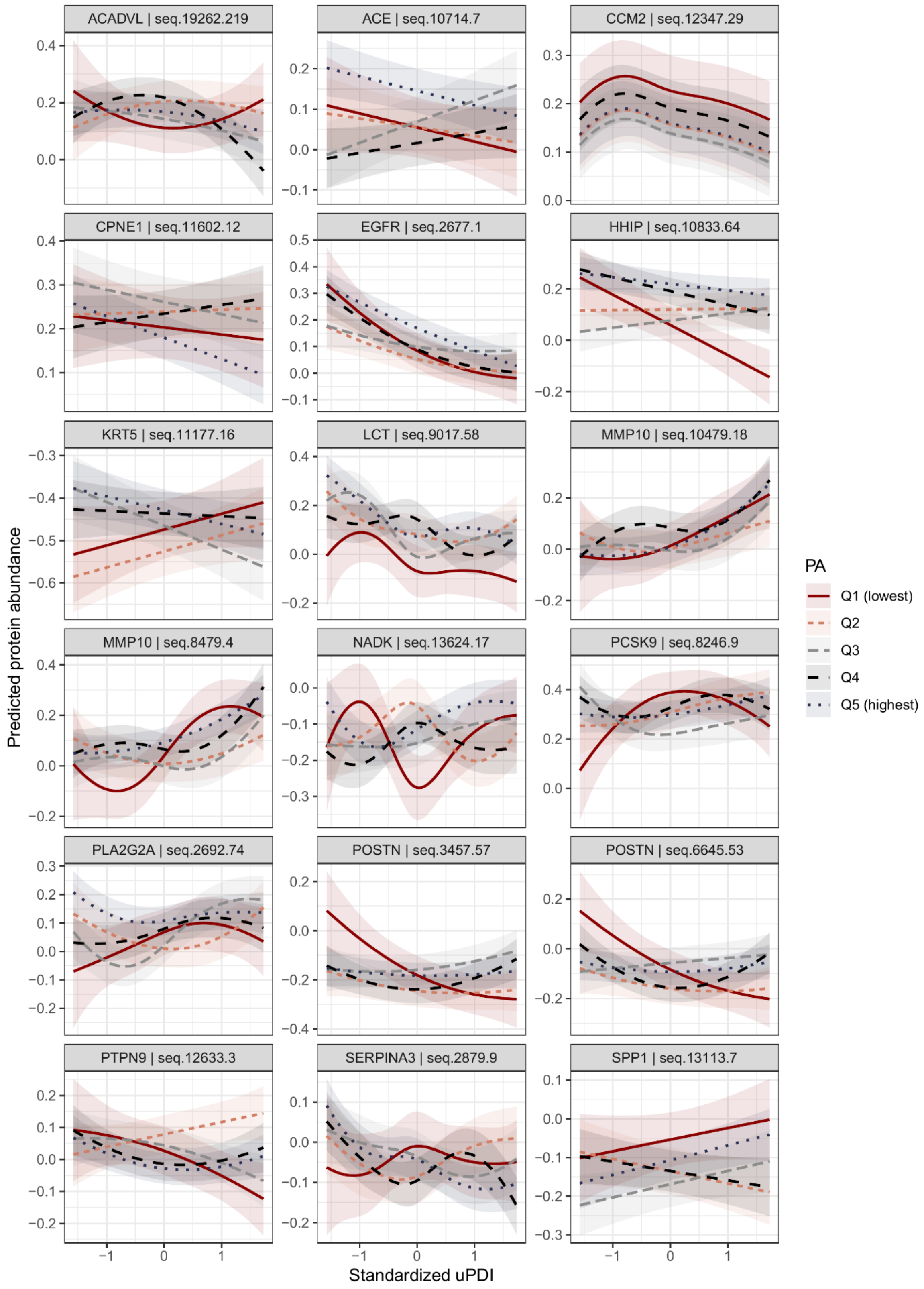

Panels show predicted protein abundance across standardized uPDI values, stratified by quintiles of physical activity levels. Models were adjusted for age, educational attainment, sex, smoking status, alcohol consumption, and country of recruitment. Age was specified as restricted cubic splines when evidence of nonlinearity was present. Lines represent physical activity quintiles (Q1–Q5), as indicated in the legend. Shaded bands indicate 95% confidence intervals. Each panel represents one protein, and y-axes are scaled separately.

Abbreviations: uPDI – unhealthful plant-based diet index, PA – physical activity, Q1- first quintile, lowest level of physical activity, Q2 – second quintile, Q3 – third quintile, Q4 - fourth quintile, Q5 - fifth quintile, highest level of physical activity

Figure S19: Top enriched functions and pathways by hPDI-derived signatures

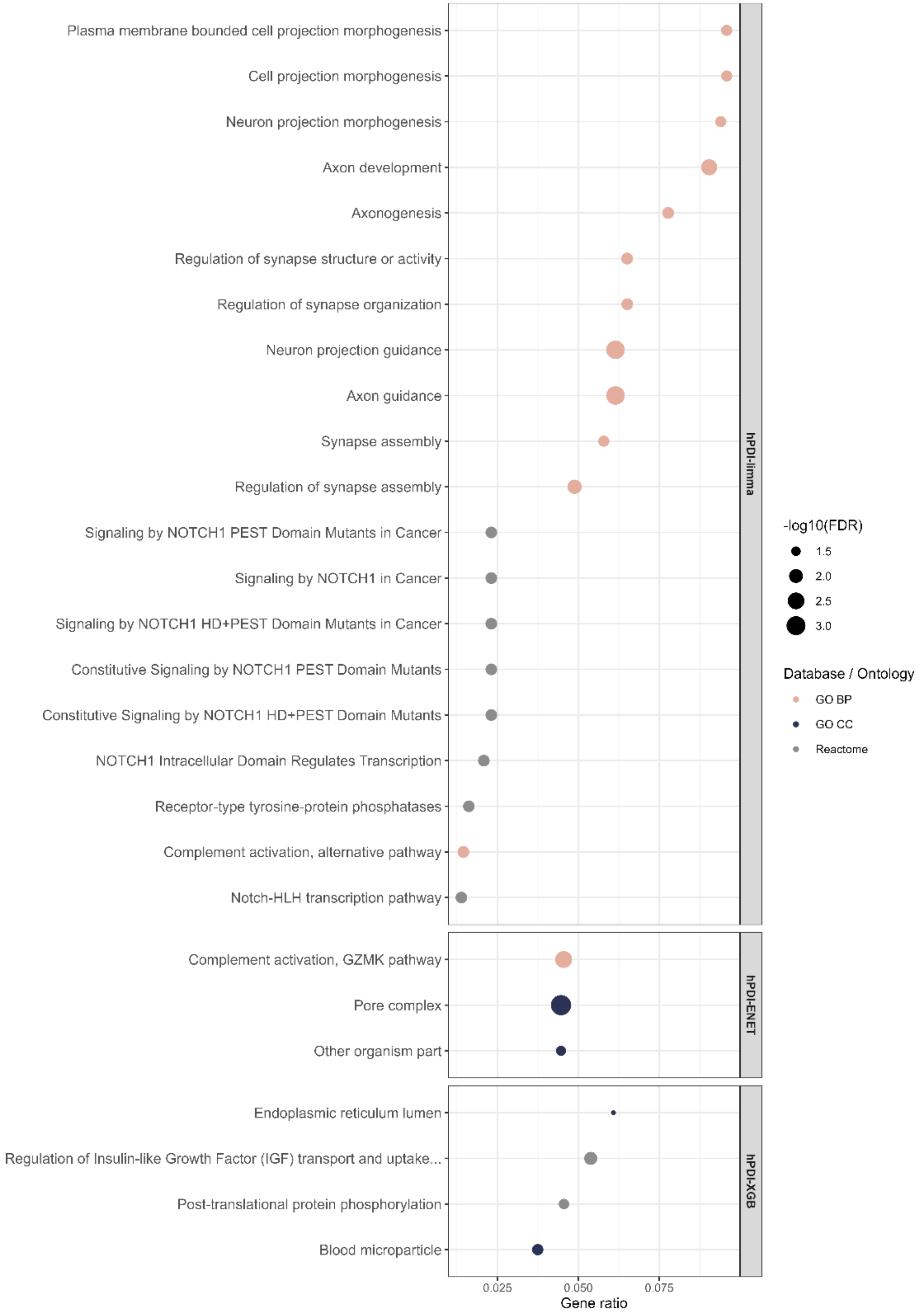

Dot plot of significantly enriched terms from Gene Ontology biological processes, cellular components, and molecular functions, KEGG, and Reactome among hPDI-related protein signatures identified by limma, ENET, and XGB. Dot size represents -log10 FDR-adjusted p-value, and dot color indicates the database or ontology source. Only terms with FDR-adjusted p values < 0.05 are shown.

Abbreviation: hPDI – healthful plant-based diet index, limma – linear models for microarray, ENET – elastic net, XGB – extreme gradient boosting, FDR - Benjamini-Hochberg false discovery rate-adjusted p value, GO BP – Gene Ontology biological processes, GO CC – Gene Ontology cellular components

Figure S20: Top enriched functions and pathways by uPDI-derived signatures

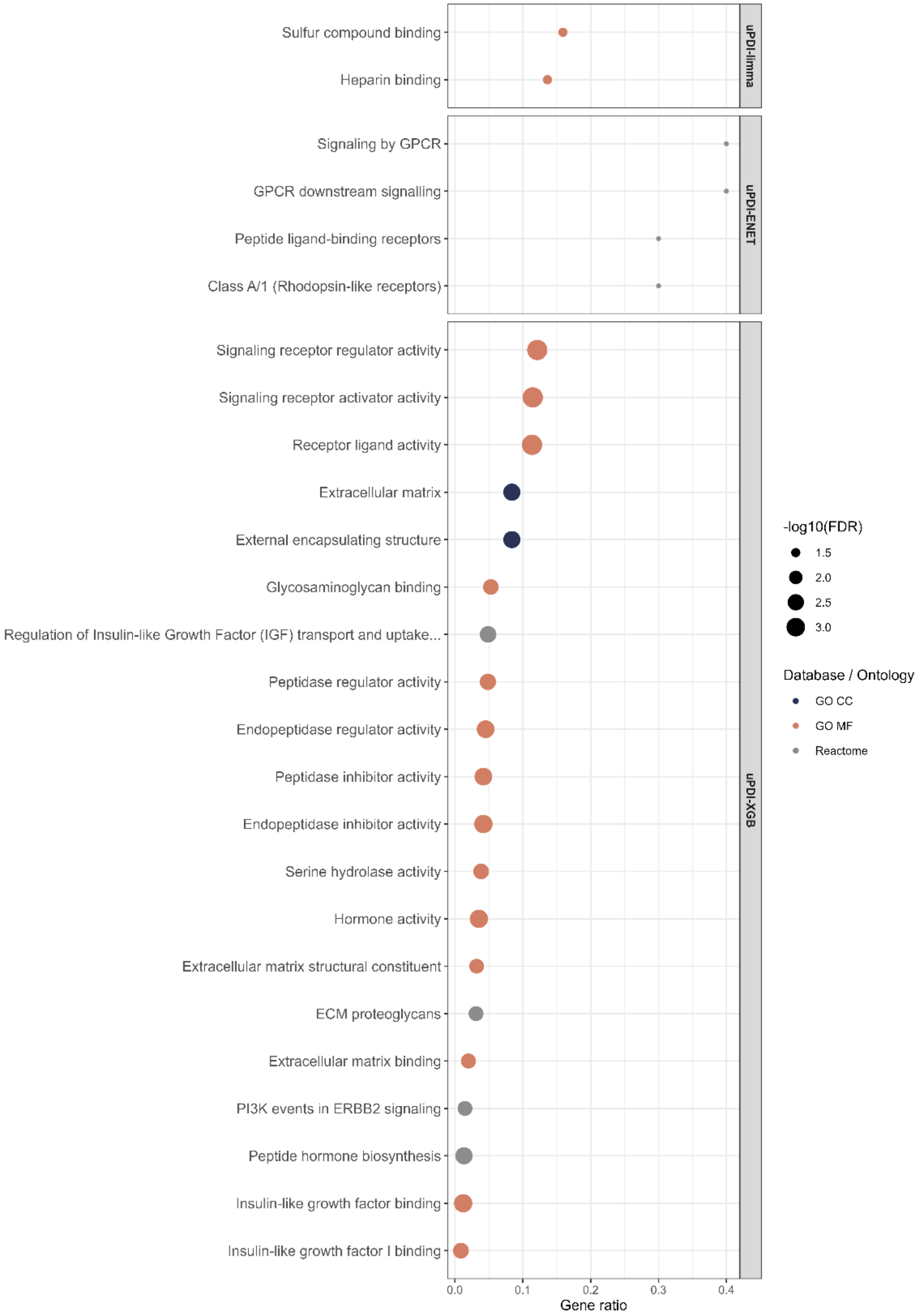

Dot plot of significantly enriched terms from Gene Ontology biological processes, cellular components, and molecular functions, KEGG, and Reactome among uPDI-related protein signatures identified by limma, ENET, and XGB. Dot size represents -log10 FDR-adjusted p-value, and dot color indicates the database or ontology source. Only terms with FDR-adjusted p values < 0.05 are shown.

Abbreviation: uPDI – unhealthful plant-based diet index, limma – linear models for microarray, ENET – elastic net, XGB – extreme gradient boosting, FDR - Benjamini-Hochberg false discovery rate-adjusted p value, GO CC – Gene Ontology cellular components, GO MF – Gene Ontology molecular functions

Figure S21: Data flow chart

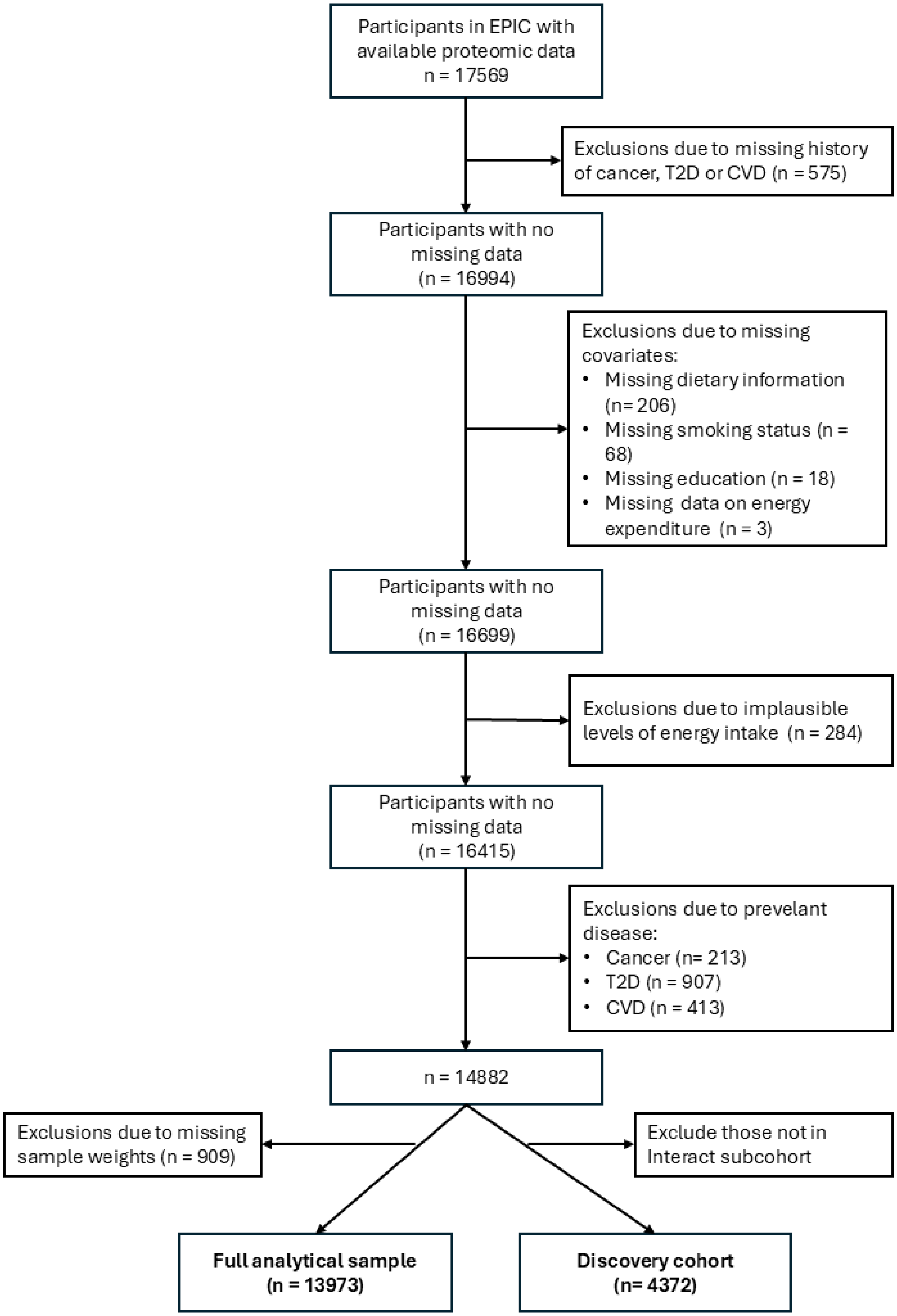

Abbreviation: n = number, T2D – type 2 diabetes, CVD – cardiovascular disease

Supplementary methods 1: Details in the derivation of dietary indices

Dietary assessment in EPIC was conducted using instruments that differed by study center. Quantitative dietary questionnaires were employed in most of Italy, Germany, Spain, and the Netherlands. On the other hand, dietary intake was assessed by semi-quantitative food frequency questionnaires in Denmark and Naples, Italy. In United Kingdom, dietary intake was assessed by semi-quantitative food frequency questionnaires and supplemented by the 7-day food records. Across these instruments, participants reported how often they consumed specific food items and the amount they consumed per occasion. The International Agency for Research on Cancer (IARC) in Lyon, France, coordinated the central processing of the dietary data. Food items from the different questionnaires were mapped to harmonized food groups across centers, including the 17 food groups used to compute the dietary indices in this study. Using reported frequencies of food intake and portion-size information, and standard portion sizes where applicable, the total daily intake for each food item was estimated in grams.

The table below describes the 17 food groups included in the computation of the dietary indices:

| | Food Groups | Examples of food items included | Scoring Scale | |
|---|---|---|---|---|
| | | | hPDI | uPDI |
| 1 | Whole grains | Muesli, oat cereals (non-sugar), bran cereals, whole meal bread, mixed bread (brown and seeded), whole meal pasta, brown rice, and other whole grains, wholewheat cereals* | Positive | Negative |
| 2 | Fruits | Blackberries, strawberries, blueberries, raspberries, cherries, grapefruit, orange, satsuma, apples, pears, bananas, mixed fruits, grapes, mango, melon, peach, pineapple, kiwi, other fruits, dried fruits such as prunes and apricots*, stewed fruits such as plums* | Positive | Negative |
| 3 | Vegetables | Garlic, leek, onion, peas, sweetcorn, broccoli, cabbage, kale, cauliflower, spinach, sprouts, mushrooms, mixed vegetables (e.g. frozen mixed vegetables or vegetable pieces too small to be counted as individual vegetable), avocado, broad beans, green beans, butternut squash, courgettes, peppers, cucumber, celery, tomatoes (fresh and tinned), root vegetables, beetroots, carrots, celery, parsnip, turnip, mixed raw salad, lettuce, water cress, other vegetables, vegetable dips such as guacamole and hummus*, vegetable side dishes with added fat dressings such as coleslaw* | Positive | Negative |

| | | | | |
|---|---|---|---|---|
| 4 | Nuts | Salted, roasted and unsalted peanuts, other nuts (e.g. cashews, almonds, pistachios), and seeds (e.g. sunflower, pumpkin, linseeds) | Positive | Negative |
| 5 | Legumes | Baked beans*, pulses (e.g. kidney beans, chickpeas, butter beans, lentils or others), vegetarian alternative to meat (e.g. vegetarian burgers, vegetarian sausages, tofu, tempeh, soya mince, Quorn)*, soy milk*, rice milk*, oat milk* | Positive | Negative |
| 6 | Tea and coffee | Coffee, instant coffee, filtered coffee, cappuccino, espresso, decaffeinated coffee (normal, instant, filter, cappuccino, espresso coffee), tea (black, green and other tea), decaffeinated tea (black, herbal rooibos), | Positive | Negative |
| 7 | Fruit juices | Orange juice, grapefruit juice, and other 100% fruit juice | Negative | Positive |
| 8 | Sugary drinks | Fizzy sugary drinks, squash, fruit smoothies, low calorie fizzy drinks, low calorie squash | Negative | Positive |
| 9 | Refined grains | Sugared oat crunch type cereals, other sweetened cereal, plain cereals, white bread (cereals may include dried fruits), white bread, baguette, bap, roll, bun, bagel, naan, garlic bread, other bread (e.g. crumpets, tortilla, wraps), white pasta, white rice, couscous, gluten free pasta, desserts, cakes and pastries (pancakes, croissant, Danish pastries, scones, fruitcakes, cakes, doughnuts, sponge puddings, cereal bars, sweet snacks, others), savoury snacks (crisps, savoury biscuits, cheese snacks, other savoury biscuits), savoury crackers (oat cakes and crispbreads including gluten free) | Negative | Positive |
| 10 | Potatoes | Mashed potatoes, fried potatoes, roast potatoes, chips, wedges, baked and boiled potatoes | Negative | Positive |
| 11 | Sweets and desserts | Chocolate biscuits, plain biscuits, sweet biscuits and cookies, chocolate bar including white, milk and dark chocolate, chocolate-covered raisins, chocolate-covered sweets, hard and soft sweets (including sugar free), soya-based desserts, soya-based yogurt, added sugar and preservatives (e.g. table sugar, honey, jam, and preserves), double and single crust pies, crumble pies, Yorkshire pudding, snack pot noodles | Negative | Positive |
| 12 | Animal fat | Dairy fat spread (normal fat or low fat), butter (normal fat or low fat) | Negative | Negative |
| 13 | Diary | Dairy-based smoothies, milk-based drinks, hot chocolate, full fat yogurt (plain), low fat yogurt (fat-free and lower fat yogurt, plain or flavoured), medium low-fat cheese (Cheese <=17.5g fat per | Negative | Negative |

| | | | | |
|---|---|---|---|---|
| | | 100 g, including hard and spreadable lower fat cheese, Cottage), High fat cheese (Cheese >17.5 g fat per 100 g, including hard cheese, soft cheese, spreadable, Blue, Feta, Mozzarella, Goats, other), milk dairy desserts (ice cream, milk puddings, milk-based desserts, cheesecake), whole milk (from cow, goat, sheep)*, semi-skimmed milk (cow, other)*, skimmed milk (cow, cholesterol lowering, powdered)*, cream (cow's milk)* | | |
| 14 | Eggs | Whole eggs (e.g. fried, boiled, poached), omelette, scrambled egg, scotch eggs, other egg dishes | Negative | Negative |
| 15 | Seafood | Fried breaded fish, battered fish, white fish (e.g. cod, haddock, fish pie), tinned tuna, other fish, oily fish (e.g. salmon, tinned salmon, herring, mackerel, sardines, fresh tuna steak), prawns, lobster, crab, other shellfish (e.g. mussels, scallops), other | Negative | Negative |
| 16 | Meat | Sausages, bacon (with and without fat), ham, liver pate or liver, ham, parma ham, salami, pastrami, cured meats, deep fried or breaded chicken or turkey, beef, pork, lamb, poultry, breaded battered chicken, other meats (e.g. duck, goose, offal) | Negative | Negative |
| 17 | Mixed foods | Pizza, gluten-free pizzas, samosa, pakora, onion bhaji | Negative | Negative |

Figure S22: Illustration of scores based on quintiles

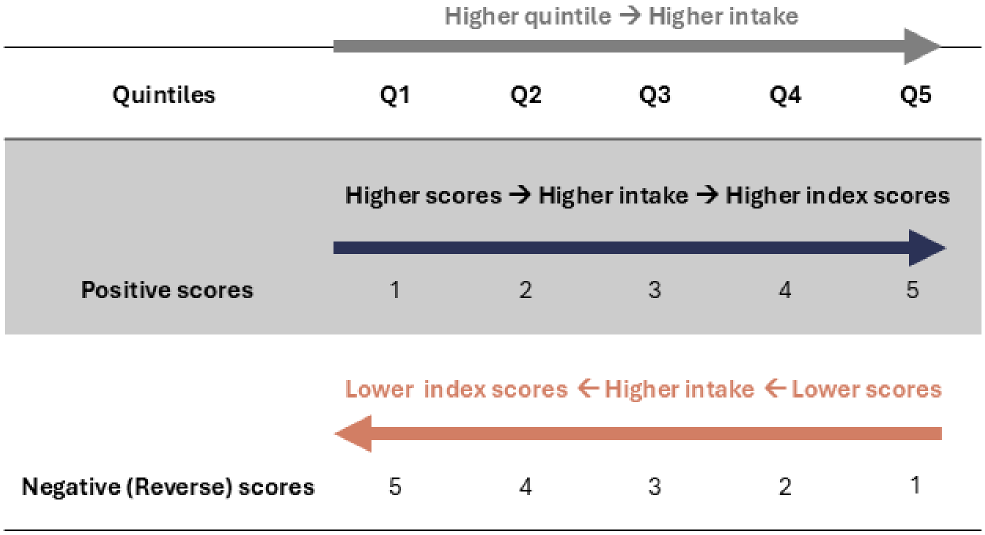

Abbreviation: Q1 – lowest quintile, lowest 20$^{th}$ of the distribution; Q2 – 20$^{th}$ to 40$^{th}$ percentile, Q3 – 40$^{th}$ to 60$^{th}$ percentile, Q4 – 60$^{th}$ to 80$^{th}$ percentile, Q5 – highest quintile, top 20$^{th}$ of the distribution

Supplementary methods 2: XGBoost hyperparameter tuning

We included all available aptamer-level protein features and the covariates, namely age at recruitment, sex, education, alcohol intake, smoking status, physical activity, and recruitment center in the XGBoost model. We did not filter aptamer-level protein features to only those found significant in linear regression models, as XGBoost allows for non-linear associations and interactions.[1] The hPDI and uPDI were standardized before model fitting and the data were split into training and testing sets using an 80:20 ratio. Hyperparameter tuning is performed only on the training set.

Firstly, we conducted a preliminary exploratory tuning to identify a suitable region of model complexity and regularization. This phase included 287 candidate hyperparameter combinations across six grids covering the following hyperparameters: maximum tree depth (max_depth), learning rate (eta), minimum child weight (min_child_weight), subsampling fraction (subsample), column subsampling fraction (colsample_bytree), L1 regularization (alpha), L2 regularization (lambda), and minimum loss reduction (gamma).[2] For each hyperparameter combination, performance was evaluated using 5-fold cross-validation in the training set. Models were trained for up to 2000 boosting rounds, with early stopping applied if the cross-validated RMSE did not improve for 50 consecutive iterations.

Secondly, the fine-tuning phase was performed around the best-performing hyperparameter region identified during preliminary tuning. This phase evaluated 18 additional candidate combinations: six combinations of max_depth and min_child_weight, nine combinations of alpha and lambda, and three values of column subsampling fraction. Performance was evaluated using 5-fold cross-validation in the training set, with models trained for up to 2000 boosting rounds and early stopping. Lastly, a final model was trained in the full training set using the best-performing hyperparameters from the fine-tuning phase and the corresponding number of boosting rounds identified during cross-validation.

References:

1. Chen, T. & Guestrin, C. XGBoost: A Scalable Tree Boosting System. in *Proceedings of the 22nd ACM SIGKDD International Conference on Knowledge Discovery and Data Mining* 785–794 (ACM, San Francisco California USA, 2016). doi:10.1145/2939672.2939785.

2. XGBoost Developers. XGBoost Parameters. https://xgboost.readthedocs.io/en/stable/parameter.html (2025).
